# Gantenerumab reduces brain amyloid-β proteoforms: Biochemical evaluation in dominantly inherited Alzheimer disease

**DOI:** 10.64898/2026.09.22.26363432

**Authors:** Soumya Mukherjee, Nicolas R. Barthélemy, Taraneh Atri, Yan Li, Daniel Yen, Yingxin He, Reid Coyle, Vitaliy Ovod, Erin E. Franklin, Chengjie Xiong, Guoqiao Wang, Jinbin Xu, Huifangjie Farsad, Catherine J. Mummery, Colin L. Masters, Raquel Sanchez-Valle, Mario Masellis, Ging-Yuek Robin Hsiung, Serge Guthier, James J. Lah, Sarah B. Berman, Erik D. Roberson, Ghulam Surti, Lawrence S. Honig, Roger Clarnette, Paolo Vitali, John Ringman, Thomas D. Bird, William S. Brooks, Nick C. Fox, Kazushi Suzuki, Sandra E. Black, Johannes Levin, Martin R. Farlow, Jasmeer Chhatwal, Neelum T. Aggarwal, Mathias Jucker, Matthew P. Frosch, Julia K. Kofler, Charles White, C. Dirk Keene, Jie Chen, Jonathan Glass, Marla Gearing, Natalie S. Ryan, David Aguillón, Charlene Supnet-Bell, Brian A. Gordon, Tammie L.S. Benzinger, John Sims, Roy Yaari, Tobias Bittner, Gregory Klein, Paul Delmar, John C Morris, Jorge Llibre-Guerra, David B. Clifford, Celeste M. Karch, Eric McDade, Richard J. Perrin, Randall J. Bateman, the Dominantly Inherited Alzheimer Network Observational Study, Dominantly Inherited Alzheimer Network-Trials Unit

## Abstract

Therapeutic strategies targeting amyloid-β (Aβ) in Alzheimer disease (AD) include antibodies directed against fibrillar amyloid plaques or soluble amyloid-β (Aβ) species, yet how these approaches reshape brain pathology remains poorly understood. Postmortem neuropathological assessment is crucial for evaluating the degree of Aβ removal, providing information and context that fluid biomarkers and positron emission tomography (PET) alone cannot, and guiding development of more effective disease modifying therapies for AD. In this study, using high resolution mass spectrometry we quantitatively measured Aβ and tau proteoforms in postmortem brain tissue homogenates to evaluate the impact of long-term treatment with anti-Aβ monoclonal antibodies gantenerumab or solanezumab in a clinical trial of dominantly inherited Alzheimer disease (DIAD). Symptomatic DIAD participants who had been treated for ∼ 4 years with anti-Aβ monoclonal antibodies (gantenerumab, n=4 or solanezumab, n=4) were compared with untreated, symptomatic DIAD mutation carriers (n=40) and with age-relevant non-DIAD controls with modest or no plaque pathology (n=18). Consistent with antemortem Aβ-PET findings in the clinical trial, gantenerumab-treated participant samples showed significantly lower levels of specific Aβ proteoforms (Aβ_42_, Aβ_40_) in the insoluble plaque associated pool (range: 60-80 % reduction); they also showed lower levels of post-translationally modified Aβ proteoform Aβ_3pGlu_ in both the insoluble and soluble pool, compared to untreated DIAD participants, but without reaching the levels found in non-DIAD controls. Samples from gantenerumab-treated participants also showed regionally lower amounts of insoluble phosphorylated tau species (pT181, pT205, pT208) and MTBR tau species (354-369, 306-317, 299-317) without apparent differences in insoluble total tau levels. In contrast, brain tissue samples from solanezumab-treated participants did not show significant differences overall in soluble or insoluble levels of Aβ and tau proteoforms in comparison to those from non-treated DIAD individuals. These findings support that amyloid targeting treatment (ATT) with gantenerumab, an antibody that targets fibrillar Aβ, can remove insoluble Aβ from the DIAD brain and also mitigate “downstream” biochemical and pathophysiological changes associated with AD.

**Key Summary:** *Question:* How do amyloid targeting treatments (ATT) affect the Alzheimer disease (AD)-related brain pathologies of soluble and aggregated amyloid-β and tau proteoforms in postmortem tissue?

*Background:* Extracellular amyloid plaques and neurofibrillary tangles (NFTs) are the primary pathological hallmarks of AD. For the past three decades, active and passive immunization against cerebral amyloid-β (Aβ) has constituted one of the primary avenues of disease-modifying intervention in AD. Recently, passive immunizations using anti-Aβ targeting antibodies have demonstrated slowing of cognitive decline in sporadic AD and possibly in dominantly inherited AD (DIAD). In these clinical trials, target engagement has been inferred from Aβ-positron emission tomography (PET) and/or biofluid biomarkers during life. However, to confirm evidence of target engagement more directly, identify which pathological proteoforms are affected, quantify the dose dependent treatment effects of these treatments, and begin to evaluate their putative downstream effects on AD pathophysiology, quantitative biochemical comparisons of amyloid plaques and tau pathology (soluble and insoluble Aβ and tau species) in postmortem brain tissue of treated and untreated AD are needed.

*Findings:* In this study, using high resolution mass spectrometry we measured the soluble and insoluble Aβ and tau proteoforms in the postmortem brain tissue homogenates from DIAN-TU-001 trial participants who were symptomatic and had been treated for ∼ 4 years with gantenerumab (n=4) or solanezumab (n=4), two antibodies that bind to insoluble and soluble Aβ, respectively. Intervals between treatment discontinuation and death ranged from 5 months to 3 years. Comparisons were made with postmortem brain tissue samples from untreated, symptomatic DIAD participants of the DIAN-Obs study (n=40) and from age-relevant, cognitively normal, non-DIAD controls with or without only modest burdens of diffuse Aβ plaques (n=18). Consistent with antemortem Aβ-PET findings in the clinical trial, gantenerumab significantly decreased specific Aβ (Aβ_42_, Aβ_40_) proteoforms in the insoluble plaque associated pool (range: 60-80% reduction) compared to the untreated DIAD participants, but without reaching the levels found in the non-DIAD controls. Samples from gantenerumab treated participants also showed significantly lower amount of post-translationally modified Aβ (Aβ_3pGlu_) and regionally lower insoluble phosphorylated tau species (pT181, pT205 and pT208), without apparent differences in insoluble total tau levels. In contrast, solanezumab treatment did not show significant effect on the soluble or insoluble levels of Aβ and tau proteoforms in comparison to samples from non-treated DIAD individuals.

*Significance:* These findings support that ATT with gantenerumab, an antibody that targets fibrillar Aβ, can remove insoluble Aβ from the DIAD brain and also mitigate “downstream” biochemical and pathophysiological changes associated with AD.

## Introduction

Alzheimer disease (AD), the most common cause of dementia, currently impacts more than 55 million people worldwide. The two main histopathological features of AD are amyloid plaques, extracellular deposits of amyloid beta (Aβ) in the parenchyma, and intraneuronal neurofibrillary tangles (NFTs) composed of hyperphosphorylated tau.^1–5^ AD is characterized by progressive neuropathological changes associated with the deposition of aggregated Aβ in the brain years before clinical symptoms emerge.^6,7^ Although most AD cases are considered sporadic and late-onset (LOAD), less than 1% of AD is dominantly inherited (DIAD), caused by pathogenic mutations in either amyloid precursor protein (*APP*), presenilin-1 (*PSEN1*), or presenilin-2 (*PSEN2*) genes. These inherited mutations promote the aggregation and extracellular deposition of Aβ peptides into amyloid plaques many years before any clinical symptoms emerge.^8,9^ This amyloid plaque formation initiates a cascade of neuroinflammation, intraneuronal tau protein hyperphosphorylation, neurofibrillary tangle (NFT) formation, and synaptic and neuronal loss,^10,11^ leading to AD dementia at a predictable and, usually, young age.^7,12^ The predictability of DIAD and its clinical and biomarker trajectories enables relatively efficient clinical trial design and the initiation of therapeutic interventions at discrete stages of the disease course.^13–15^ Although there are regional variabilities in amyloid pathology between LOAD and DIAD, with DIAD having relatively greater regional burdens,^16^ neuroimaging and biofluid biomarkers suggest that these two forms of AD share a common ‘downstream’ pathophysiology.^17,18^ Thus, findings from studies of either may be relevant to both, and trials in DIAD have predicted outcomes in LOAD trials.^19^

In clinical trials of both DIAD and LOAD, several amyloid targeting treatments (ATT), target different forms of Aβ peptides with monoclonal antibodies, and those that remove Aβ aggregates^20–25^ have been shown to reduce plaque burden and slow the rate of cognitive decline. In the phase 2/3 placebo-controlled, double-blind, randomized clinical trial in the Dominantly Inherited Alzheimer Network Trial Unit (DIAN-TU-001, NCT04623242)^13^, two ATT monoclonal antibodies (gantenerumab and solanezumab) were investigated. Gantenerumab, which targets fibrillar Aβ, initiates plaque removal via fragment crystallizable (Fc) γ-receptor-mediated activation of microglial phagocytosis.^26^ In contrast, solanezumab targets the clearance of soluble Aβ from the brain by binding to the soluble monomeric Aβ.^27^ Findings from the DIAN-TU-001 trial and the subsequent gantenerumab open label extension (OLE) showed gantenerumab-mediated reduction of amyloid burden, inferred by Aβ positron emission tomography (PET) signal reduction and/or changes in CSF levels of Aβ42/40, p-tau181, and p-tau217.^14,28–30^ Nevertheless, how gantenerumab and solanezumab treatments might alter different Aβ and tau proteoforms and other AD-related pathologic features and in the DIAD brain remains unknown.

Postmortem neuropathological studies of patients who received anti-amyloid monoclonal antibodies antemortem are key to understanding and correlating the ATT-associated pathophysiological and biochemical changes in the brain with neuroimaging and biofluid biomarker modalities.^31–34^ Additionally, they allow for investigation into other downstream pathological brain changes such as tauopathy burden, neuroinflammation, and spatial omics changes for which antemortem biomarkers have not yet been developed.^34,35^ A postmortem immunohistochemical analysis of DIAN-TU-001 participants and DIAD controls has confirmed a dose-dependent reduction of amyloid plaque burden with gantenerumab treatment ^27^ but how gantenerumab and solanezumab treatments might alter the aggregation states and different proteoforms of Aβ and tau in the DIAD brain remains incompletely understood.

In this study, we examined the impact of treatment with ATTs in DIAN-TU-001 participant brain donors who received treatment with gantenerumab (during the trial and the gantenerumab open label extension, or only during the trial [n=4]), or solanezumab (n=4), by comparing them with untreated DIAN Observational Study (DIAN-Obs) participant brain donors (n=40) and age-relevant non-DIAD cognitively normal controls with no or minimal amyloid plaque pathology (n=18). Using high resolution liquid chromatography mass spectrometry (LC-MS), we quantitatively measured the biochemical pools of soluble and insoluble Aβ and tau proteoforms derived from brain tissue (primarily grey matter) homogenates across multiple brain regions.^36,37^ Our investigation into the effect of each drug on Aβ and tau constituting the two pathological hallmarks of AD, amyloid plaques and tau tangles, in postmortem brain tissue provides support for the continued development of anti-amyloid monoclonal antibodies directed towards insoluble Aβ in ameliorating AD neuropathological changes.

## Methods

### Trial Design and Participants

The DIAN-TU-001 study was approved by the Washington University Human Research Protection Office and local institutional review boards as described previously^15^. Some of the data used in the preparation of this article were obtained from the Dominantly Inherited Alzheimer Network Trials Unit (DIAN-TU). The DIAN-TU was launched in 2011 as a public-private partnership to implement effective, safe and efficient clinical trials that have the highest likelihood of success in advancing overall treatments as well as scientific understanding of dominantly inherited Alzheimer disease. For up-to-date information, see https://dian.wustl.edu/.

Participants with DIAD included in this study were either enrolled in the Dominantly Inherited Alzheimer Network Observational Study (DIAN-Obs, n= 40) or DIAN-TU-001 (n= 8). The non-DIAD control brain samples (NC, n= 18) were provided by Emory University (Table 1 and Supplementary Table 1).

**Table 1.** Participant characteristics of the DIAD observational mutation carriers (DIAN Obs MC), Dominantly Inherited Alzheimer Network trial unit (DIAN-TU-001) trial participants, and non-DIAD cognitively normal controls included in this study.

| Characteristics | DIAN Obs MC<br>(n = 40) | Gantenerumab<br>(n= 4) | Solanezumab<br>(n = 4) | Non-DIAD Controls<br>(n = 18) |
| --- | --- | --- | --- | --- |
| Age at death (years ± SD) | 53.0 ± 9.1 | 54.3 ± 7.9 | 50.8 ± 12.1 | 51.7 ± 9.5 |
| Female, n (%) | 16 (40) | 0 (0) | 2 (50) | 8 (44.4) |
| *APP* mutation carriers, n (%) | 6 (15) | 1 (25) | 0 (0) | N/A |
| *PSEN1* mutation carriers, n (%) | 32 (80) | 3 (75) | 4 (100) | N/A |
| *PSEN2* mutation carriers, n (%) | 2 (5) | 0 (0) | 0 (0) | N/A |
| *APOE-ε4* carriers, n (%)<sup>e</sup> | 10 (40)<sup>a</sup> | 3 (75) | 0 (0) | 6 (38)<sup>d</sup> |
| CDR<sup>®</sup> Baseline (mean ± SD) | 1.22 ± 0.75<sup>a</sup> | 0.63 ± 0.25 | 1.00 ± 0.00 | N/A |
| CDR<sup>®</sup> Baseline (0/0.5/1/2/3) | (0/7/11/5/2)<sup>a</sup> | 0/3/1/0/0 | 0/0/4/0/0 | N/A |
| CDR<sup>®</sup> SB Baseline (mean ± SD) | 6.88 ± 4.55<sup>a</sup> | 2.50 ± 1.83 | 5.75 ± 0.96 | N/A |
| MMSE Baseline (mean ± SD) | 16.84 ± 7.30<sup>a</sup> | 24.75 ± 4.43 | 18.00 ± 0.82 | N/A |
| CDR<sup>®</sup> last visit (0/0.5/1/2/3) | (0/0/6/14/5)<sup>a</sup> | 0/0/1/1/2 | 0/0/0/2/2 | N/A |
| Expiration CDR (0/0.5/1/2/3) | (0/0/0/0/38)<sup>b</sup> | (0/0/0/1/3) | (0/0/0/1/3) | N/A |
| MMSE last visit (mean ± SD) | 11.92 (5.57)<sup>a</sup> | 9.25 (3.40) | 3.75 (5.68) | N/A |
| Thal Phase (0/1/2/3/4/5) | (0/0/0/0/0/24)<sup>c</sup> | (0/0/0/1/0/3) | (0/0/0/0/0/4) | (10/3/4/0/0/0) |
| Braak stage (0/I/II/III/IV/V/VI) | (0/0/0/0/0/1/23)<sup>c</sup> | (0/0/0/0/0/4) | (0/0/0/0/1/3) | (0/6/4/0/0/0/0) |

<sup>a</sup> Indicates only participants (n =25) for which the associated data is available
<sup>b</sup> Indicates only participants (n=38) for which the associated data is available
<sup>c</sup> Indicates only participants (n=24) for which the associated data is available
<sup>d</sup> Indicates only participants (n=17) for which the associated data is available
<sup>e</sup> Participants with at least one *APOE ε4* allele
*APOE*, Apolipoprotein E genotype; *APP*, Aβ precursor protein; CDR<sup>®</sup>, Clinical Dementia Rating<sup>®</sup>; MMSE, Mini-Mental State examination; *PSEN1*, presenilin-1; *PSEN2*, presenilin-2; SD, standard deviation
N/A Data not available

### Cohort demographics

For the present analysis, we included DIAN-TU-001 participants enrolled in the gantenerumab (n=4) or solanezumab arms (n=4) with Clinical Dementia Rating^®^ [CDR^®^] scores of 0.5 or 1 at baseline. Three out of four of the gantenerumab-treated participants continued to receive treatment during the gantenerumab open label extension period. All eight progressed to moderate (CDR 2 [n=2]) or severe AD dementia (CDR 3 [n=6]) before expiration. The durations, cumulative doses, and intervals between last dose and expiration were variable across participants for both ATTs (Table 2). For gantenerumab, exposures ranged from 2 to 6 years, cumulative doses ranged from 5850 mg to 48420 mg, and last dose to expiration intervals ranged from 5 months to 2 years. For solanezumab, exposures ranged from 1 to 4 years, cumulative doses ranged from 6400 mg to 48400 mg, and last dose to expiration intervals ranged from 1 year to 3 years.

**Table 2.** Anti-Aβ antibody treated patient enrolled in DIAN-TU-001 demographics.

| DIAN-TU-001 Participant | Sex (M/F) | *APOE* Genotype | Mutation Type | Age at baseline | Age of Onset | CDR<sup>®</sup> at baseline | EYO at baseline | Drug start EYO | Drug | Total drug received (mg) | Treatment end EYO | Disease Duration | EYO at death | Treatment end to Death (years) | Age at death | CDR<sup>®</sup> at death |
| --- | --- | --- | --- | --- | --- | --- | --- | --- | --- | --- | --- | --- | --- | --- | --- | --- |
| 1 | M | $\epsilon 3\epsilon 4$ | *PSEN1* | 40-45 | 42-47 | 0.5 | -3.0 | -3.0 | Gant | 27045 | 1.9 | 2.9 | 3.8 | 1.8 | 48-53 | 3 |
| 2 | M | $\epsilon 3\epsilon 4$ | *APP* | 45-50 | 47-52 | 0.5 | 0.5 | 0.5 | Gant | 5850 | 2.4 | 4.2 | 4.6 | 2.2 | 50-55 | 2<sup>a</sup> |
| 3 | M | $\epsilon 3\epsilon 4$ | *PSEN1* | 55-60 | 60-65 | 0.5 | -1.2 | -1.2 | Gant | 48420 | 4.0 | 5.4 | 6.1 | 2.2 | 66-71 | 3 |
| 4 | M | $\epsilon 2\epsilon 3$ | *PSEN1* | 41-45 | 36-41 | 1 | 3.2 | 3.2 | Gant | 35400 | 8.8 | 8.7 | 9.2 | 0.4 | 46-51 | 3 |
| 5 | F | $\epsilon 3\epsilon 3$ | *PSEN1* | 35-40 | 36-41 | 1 | -1.9 | -1.9 | Sola | 48400 | 2.1 | 3.0 | 3.5 | 1.4 | 36-41 | 3 |
| 6 | M | $\epsilon 3\epsilon 3$ | *PSEN1* | 57-61 | 51-56 | 1 | 7.3 | 7.3 | Sola | 20400 | 11.2 | 12.0 | 12.7 | 1.5 | 61-66 | 2<sup>a</sup> |
| 7 | M | $\epsilon 2\epsilon 3$ | *PSEN1* | 50-55 | 46-50 | 1 | 7.4 | 7.4 | Sola | 7200 | 8.8 | 10.6 | 11.0 | 2.2 | 56-60 | 3 |
| 8 | F | $\epsilon 3\epsilon 3$ | *PSEN1* | 34-39 | 25-30 | 1 | 7.7 | 7.7 | Sola | 6400 | 8.9 | 11.0 | 11.8 | 2.9 | 36-40 | 3 |

<sup>a</sup> No expiration CDR was available for these participants and represents the last measured CDR value
EYO, estimated year of onset; *Gant*, Gantenerumab; *Sola*, Solanezumab; *APOE*, Apolipoprotein E genotype; *APP*, Aβ precursor protein; CDR<sup>®</sup>, Clinical Dementia Rating<sup>®</sup>; *PSEN1*, presenilin-1

The DIAN-Obs Study participant brain donors, with pathogenic variants in *PSEN1* (80%), *PSEN2* (5%), and *APP* (15%), were included in this study (n = 40) to represent untreated DIAD carriers with ages and a pathogenic variant distribution resembling those of the DIAN-TU-001 participant brain donors. All progressed to severe AD dementia (CDR 3) before expiration (Table 1).

The DIAD non-carriers (NC) brain donors were included in this study (n=18) to represent an age-relevant control group without cognitive impairment and with modest amounts or no neuropathologic evidence of Aβ aggregates (Thal Aβ phases 0 [n=10], 1 [n=3], or 2 [n=4], or unknown [n=1]) at autopsy (Table 1).

## Materials

All LC-MS grade solvents including acetonitrile (ACN), formic acid (FA), trifluoracetic acid (TFA), methanol and NH_4_OH were purchased from Millipore-Sigma. IGEPAL (branched octylphenoxy poly(ethyleneoxy)ethanol), tris-buffer saline (TBS), phosphate buffer saline (PBS), guanidine hydrochloride, NaCl, urea, thiourea, N, N-Bis(2-hydroxyethyl)glycine (Bicine), 3-[(3-Cholamidopropyl)dimethylammonio]-1-propanesulfonate hydrate (CHAPS), triethylammonium bicarbonate (TEABC), hydrogen peroxide (H_2_O_2_), iodoacetamide (IAA), tri-ethyl ammonium bicarbonate buffer, NaCl, Na_2_CO_3_, ethylenediaminetetraacetic acid (EDTA), ethylene glycol-bis(β-aminoethyl ether)-N,N,N’,N’-tetraacetic acid (EGTA), N-Lauroylsarcosine sodium and DL-dithiothreitol (DTT) were purchased from Millipore-Sigma or ThermoFischer Scientific. Phosphatase inhibitor cocktail 3, cOmplete™ protease inhibitor cocktail, human serum albumin (HSA) and bovine serum albumin (BSA) were purchased from Millipore-Sigma. Bond-Breaker^TM^ tris(2-carboxyethyl)phosphine (TCEP) Solution, neutral pH and Dynabeads™ M-270 Epoxy were purchased from ThermoFischer Scientific. MS grade metalloprotease Lys-N was from Immunoprecise, MS grade enzyme Lys-C was from ThermoFischer Scientific and sequencing grade trypsin was from Promega. Oasis HLB µElution 96 well-plates were purchased from Waters.

### Human Brain tissue homogenization and biochemical fractionation

Frozen brain tissue samples were processed for Aβ and tau extraction as described previously (Figure 1).^36–38^ Representative blocks of frozen tissue, maintained at −80℃, were prepared by the DIAN Neuropathology Core from eight major brain regions: inferior parietal lobule, dorsolateral prefrontal cortex, superior temporal gyrus, striatum (caudate and putamen), cerebellum, thalamus, pons, and occipital cortex. Cortical and cerebellar blocks contained >50% gray matter. Each tissue piece was individually embedded in optimal cutting temperature (OCT) compound, oriented to expose the desired anatomical plane for cryosectioning, and sectioned at 100 µm using a cryostat. Gray matter was carefully isolated from each 100 µm section under visual guidance to maintain anatomical consistency across regions. Approximately 250 mg of gray matter was collected from each block for mass spectrometry analysis of insoluble Aβ and tau, and 10 mg for analysis of soluble Aβ and tau. All the tissue homogenization and biochemical extraction was performed by individuals blinded to disease, mutation and treatment status.

**Figure 1:**
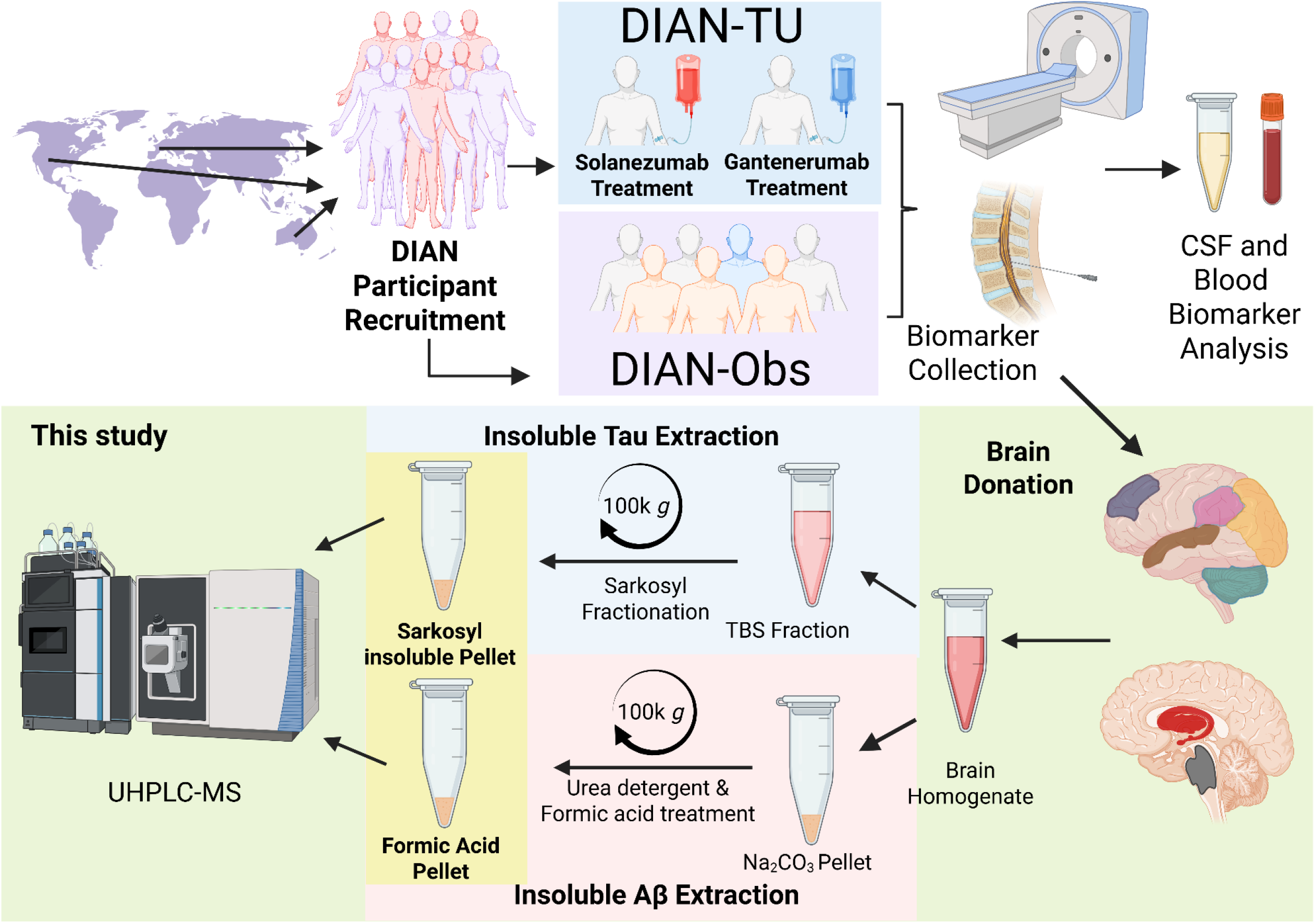
Workflow for the biochemical extraction of neuropathological proteins from DIAN-TU participant brains. Schematic workflow depicting the quantitative biochemical analyses of the levels of insoluble Aβ and tau proteoforms from frozen postmortem brain tissue samples donated by individuals with DIAD mutations who were recruited as DIAN participants, treated with anti-Aβ targeting monoclonal antibodies (DIAN-TU-001), and later died with AD dementia. Clinical, imaging and biofluid biomarkers for the recruited participants were also collected during life. In this study (green region), samples were processed and analyzed using from ultra-high performance liquid chromatography-mass spectroscopy (UHPLC-MS). The workflow for DIAN-Obs participants included in this study was similar but lacked the gantenerumab/solanezumab treatment step. The workflow for non-DIAD control brain donors prior to brain donation is not depicted. The brain regions investigated in this study are highlighted: dorsolateral prefrontal cortex (blue), superior temporal gyrus (brown), parietal cortex (pink), occipital cortex (orange), striatum (caudate and putamen) (red), pons (grey) and cerebellum (green). Created with Biorender.comx2

Preparation of soluble brain homogenate: 5-10 mg brain aliquots were sonicated (Fisherbrand sonicator [model 120 Sonic Dismembrator FB120], for 15 seconds with 1 sec on and 1 sec off pulses using 35% intensity) at 4℃ in 10 µL homogenization buffer (1% IGEPAL, phosphate inhibitors, phosphate inhibitors in 1X PBS pH 7.4, 5 mM guanidine hydrochloride) per mg of brain tissue. Homogenates were then centrifuged at 11,000 *g* for 20 minutes at 4℃. The pellet was discarded, and the supernatant was aliquoted into new Axygen tubes and kept at −80 ℃ until use.

Preparation of insoluble Aβ using sequential biochemical fractionation (as previously described, with some modifications):^36,39^ Briefly, 250 mg grey matter aliquots were sonicated (Fisherbrand sonicator [model 120 Sonic Dismembrator FB120], for 15 seconds with 1 sec on and 1 sec off pulses using 35% intensity) at 4℃ in 810 µL tris-buffered saline (TBS, 50 mM tris-HCl, 150 mM NaCl, 10 mM EDTA, 10 mM EGTA, pH7.4), containing protease inhibitors and phosphatase inhibitors (1:3, tissue:buffer). Homogenates were centrifuged at 11,000 *g* for 20 minutes at 4℃. The supernatant (TBS fraction) was aliquoted for sarkosyl insoluble tau extraction (described in next paragraph). The resulting pellet was sequentially extracted for fibrillar plaque Aβ. First, the tissue pellet was resuspended in 100 mM Na_2_CO_3_ pH 11 and incubated for 20 minutes on ice before ultracentrifugation. The samples were transferred to ultracentrifuge tubes (Beckman Coulter) and centrifuged (Optima MAX-XP from Beckman Coulter) at 100,000 *g* for 30 minutes at 4℃. The supernatant (Na_2_CO_3_ fraction containing peripheral membrane and vesicular material) was transferred into a 1.7 mL Eppendorf tube (Axygen, MCT-175-L-C). The resulting tissue pellet was further resuspended with urea-detergent buffer (7M urea, 2 M thiourea, 4% CHAPS, 30 mM bicine, pH 8.5) and spun at 100,000 *g* for 30 minutes at 4℃. The lipid layer was removed carefully, and the supernatant was aspirated out (urea-detergent fraction containing membranous proteins). The residual pellet was washed with TBS buffer twice, dried and then was finally incubated with 99% FA at 1:1 ratio (tissue:formic acid, weight/volume) for 2 hours at room temperature in a fume hood and then centrifuged at 13,200 g for 15 minutes at 4℃. The formic acid (FA) fraction (plaque Aβ enriched fraction) was aliquoted into 10 µL portions and snap frozen in liquid N_2_, freeze-dried in a lyophilizer, and stored at −80℃ until use.

Preparation of insoluble tau (as previously described):^38,40^ The crude TBS supernatant (from previous section) was treated with N-laurylsarcosinate (1% [weight/volume] final concentration), incubated on ice for 60 minutes, then ultracentrifuged (Optima MAX-XP from Beckman Coulter) at 100,000 *g* for 1 hour at 4℃ using ultracentrifuge tubes (Beckman Coulter). The sarkosyl-insoluble pellet was washed with washing buffer (TBS, 50 mM tris-HCl, 150 mM NaCl, pH 7.4), dried and finally resuspended in 100 µL of 20 mM Tris-HCl, pH 7.4 and stored at −80°C until further use. The sarkosyl insoluble pellet was aliquoted in 10 µL portions and treated with 99% FA for 2 hours at 22°C in the thermomixer (Eppendorf ThermoMixer C) at 300 rpm. These FA treated sarkosyl insoluble tau pellets were stored at −80°C until further use.

### Soluble and insoluble Aβ quantitation using immunoprecipitation mass spectrometry

Using a mid-domain anti-Aβ monoclonal antibody (HJ5.1) (mid-domain region epitope spanning residues 13-28), we immunoprecipitated and investigated Aβ proteoforms using targeted mass spectrometry (IP-MS). We quantified enzymatically digested Aβ peptides across the N-terminus, mid-domain and C-terminus of the Aβ sequence (Figure 2A). For the soluble Aβ analysis, we IP Aβ using HJ5.1 antibody as previously described with some modifications.^40,41^ To each soluble brain homogenate (40 µL, derived from 4 mg brain tissue), 0.01% HSA solution was added along 20 µL cocktail of ^15^N labeled recombinant internal standards (0.1 ng/µL Aβ_1-42_, 0.01 ng/µL Aβ_1-40_, 0.01 ng/µL Aβ_1-43_ and 0.01 ng/µL Aβ_1-38_ prepared in 0.1% NH_4_OH/20% ACN). Each sample was spiked with 50 µL of HJ5.1 coupled DynaBeads (25 µg antibody/mg beads per IP) and 25 µL of 1% IGEPAL, 1X protease inhibitor, 5 mM guanidine hydrochloride in PBS pH 7.4. The IP and bead washing steps were performed on a KingFisher (ThermoFisher Scientific) automated format, Aβ peptides were eluted off the beads using neat FA (99%) and lyophilized to dryness. The samples were resuspended in 100 mM TEABC buffer, pH 8 and 50 ng metalloprotease Lys-N (Immunoprecise) was added for enzymatic digestion for 18 hours at room temperature. The enzymatically digested samples were desalted using an Oasis HLB µElution plate (Waters) using the manufacturer’s protocol, and lyophilized. The digested peptides were incubated with 3% H_2_O_2_, 3% FA for 18 hours at 4 ℃ to oxidize the Aβ peptides containing methionine residues. The oxidized samples were desalted again using an Oasis HLB µElution plate (Waters) and lyophilized for MS analysis.

**Figure 2.**
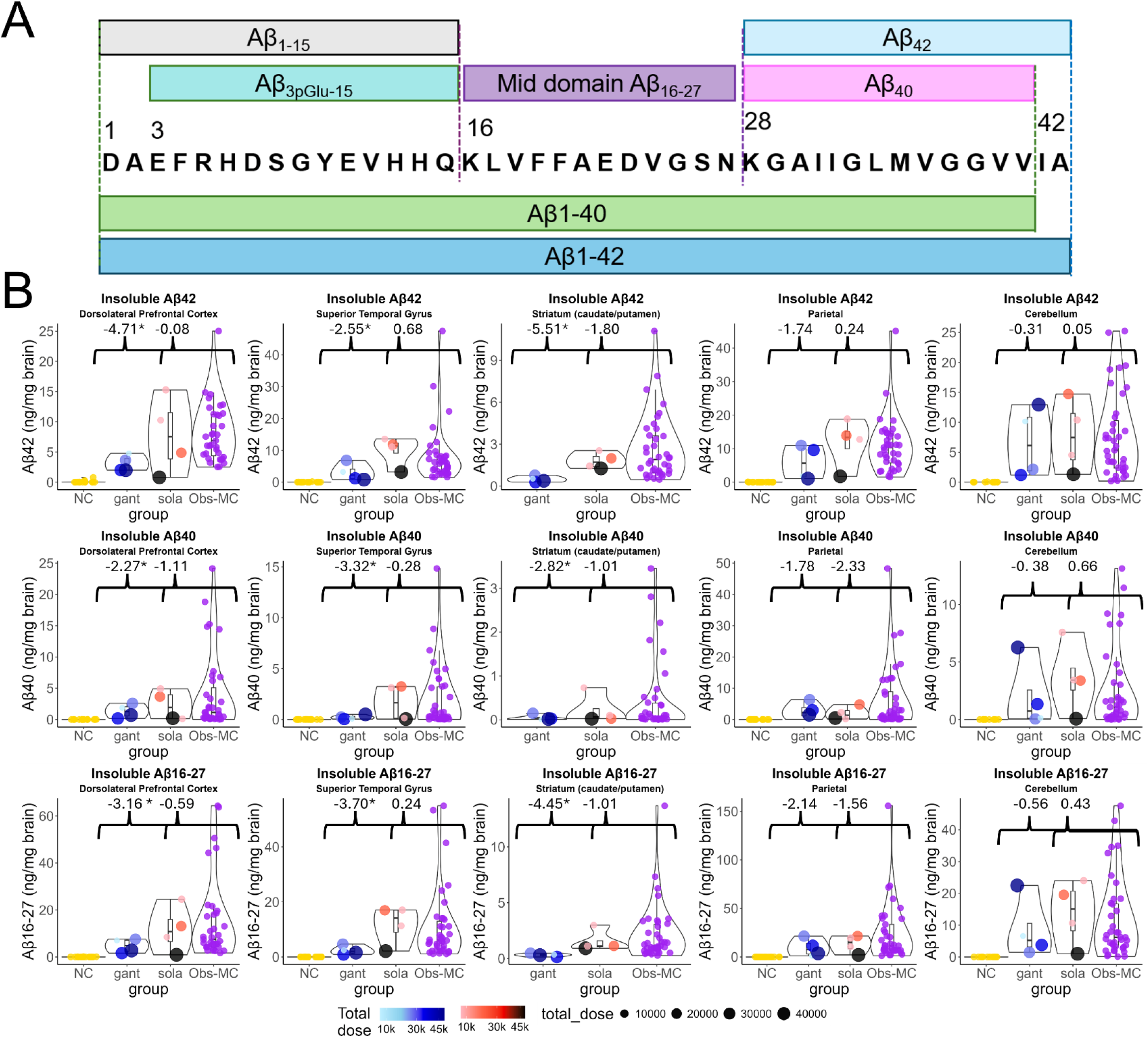
Dose dependent reduction of insoluble Aβ_42_ and Aβ_40_ brain levels in symptomatic DIAD mutation carriers treated with gantenerumab. (A) Schematic representation of canonical Aβ sequence. Aβ_1-40_ and Aβ_1-42_, highlighted in green and blue, respectively, are produced by the activity of BACE and γ-secretase in the amyloidogenic pathway from APP. The current study focuses on the Lys-N enzyme cleaved peptides of Aβ that were quantitatively measured using high resolution mass spectrometry following a immunoprecipitation (IP) using mid-domain epitope binding (residues spanning 13-28) monoclonal HJ5.1 antibody; Aβ_1-15_, Aβ_pyroGlu3-15_, Aβ_42_ (Aβ_28-42_), Aβ_40_ (Aβ_28-40_) and Aβ_mid-domain_ (Aβ_16-27_) highlighted in gray, teal, pink, yellow and purple, respectively. (B) Scatter plots of quantitative measures of the insoluble (top row) Aβ_42_ (ng/mg brain), (middle row) Aβ_40_ (ng/mg brain) and (bottom row) Aβ_mid-domain_ (ng/mg brain) in different brain regions (dorsolateral prefrontal cortex, superior temporal gyrus, striatum, parietal cortex, and cerebellum) from the postmortem brains of age-relevant, cognitively normal non-DIAD controls (NC, n =18), DIAD mutation carriers treated with gantenerumab (gant, n=4) or solanezumab (sola, n=4) and untreated observational DIAD mutation carriers (DIAN Obs-MC, n=40). The total cumulative dose (mg) of the drug received in the clinical trial for gant (blue) and sola (red) is highlighted in color gradient for each participant. Asterisks denote FDR adjusted *p*-values < 0.05 (*) associated with Welch two-sample t-tests comparing Aβ isoforms between the Obs-MC group and the gantenerumab arm or the solanezumab arm.

For insoluble Aβ analysis, the lyophilized formic acid (FA) brain fraction aliquots were reconstituted in 1 M tris-HCl, pH 8.8 buffer and diluted to 500 µL using 0.01% HSA. Insoluble Aβ was immunoprecipitated from these samples using HJ5.1 antibody as described above.

Both the soluble and insoluble Aβ peptides from brain homogenates were independently reconstituted in 2% ACN/2% FA containing 20 fmol/µL BSA tryptic peptides for mass spectrometry (MS) analysis. The peptides were analyzed on a nano-Acquity LC (Waters Corporation, Milford, MA) coupled to an Orbitrap Fusion (ThermoFisher Scientific, San Jose, CA, USA). Peptides from the soluble Aβ IP were directly loaded in 2% buffer B (0.1% FA in acetonitrile) onto an HSS T3 75 μm × 100 μm, 1.8 μm C18 column (Waters) that was heated to 65 ℃ and were separated with a flow rate of 0.4 µL/min using buffer A (0.1% FA in water) and buffer B. The Aβ peptides were eluted off the column with a gradient of 2-5% buffer B for 7 min, 5-30% for another 20 min before ramping to 80% buffer B to clean the column for the next 2 min. The peptides from the insoluble Aβ IP were loaded directly in 0.5% buffer B and separated using the same LC-MS configuration as described above and the gradient used as follows: 0.5-4% buffer B for 7 min, 4-8% buffer B for next 7 min, 8-20% buffer B for another 7 min, 20-35% buffer B for 5 min before ramping to 90% buffer B and cleaning the column for the next 2 min. The Orbitrap Fusion was equipped with a Nanospray Flex electrospray ion source (Thermo Fisher Scientific) and operated in positive ion mode. Proteolytic peptides were sprayed from a 10 µm SilicaTip emitter (New Objective, Woburn, MA, USA) into the ion source (spray voltage = 2200 V, ion source temperature 275 ℃), peptides were targeted and isolated in the quadrupole and fragmented by higher-energy collisional dissociation (HCD) and the ion fragments were detected in the Orbitrap (resolution of 30,000; mass range 150-1500 *m/z*). The raw MS data were imported into Skyline (version 20.2 MacCoss Lab, Department of Genome Sciences, University of Washington) with formula annotations of the targeted peptides. Five Aβ peptides (Aβ_1-15_, Aβ_3pGlu-15_, Aβ_16-27_, Aβ_28-42_, Aβ_28-40_) were quantified by comparison with the corresponding isotopomer signals from their respective ^15^N internal standard (^15^N Aβ_1-15_ peak area was used to quantitate Aβ_3pGlu-15_).^36,37,41^

### Soluble and insoluble tau quantitation using immunoprecipitation mass spectrometry

For the soluble tau analysis, we immunoprecipitated tau using tau1, HJ8.5 and HJ8.7 antibody mixture as previously described with some modifications.^40,42,43^ To each soluble brain homogenate (30 µL, ∼ 3 mg brain tissue), 0.01% HSA solution was added along with recombinant uniformly labeled^15^N tau 2N4R (2.5 ng, gift from Dr. Guy Lippens, Lille, France) and ^15^N tau 0N3R (2.5 ng, Promise Proteomics, Grenoble, France) as protein internal standards. Tau from each sample was immunoprecipitated using a 20 µL cocktail of conjugated beads (mix of 50% slurry of each tau antibody conjugated Sepharose beads containing 3 µg antibody/mg beads) and incubating them with rotation for 18 hours at 4℃. Following incubation, the samples were washed three times with 25 mM TEABC buffer. The proteins were then eluted off the Sepharose beads using 99 % FA and the eluent was lyophilized. The lyophilized samples were resuspended in 25 mM TEABC and reduced using 5 µL of 100 mM DTT for 30 min at 65 ℃. The reduced sample was alkylated using 5 µL of 100 mM IAA for 45 minutes in the dark at 25 ℃. The proteins were digested overnight using 0.4 µg trypsin at 37 ℃. The digested samples (soluble tau) were desalted using the Oasis µElution HLB plate (Waters) according to manufacturer’s protocol. Prior to desalting, the samples were spiked with stable isotope labeled synthetic respective phosphorylated and unphosphorylated peptides AQUA peptide standards (50 fmol non-phosphorylated peptides and 5 fmol AQUA phosphorylated peptides). The digested peptides were desalted using the Oasis µElution HLB plate (Waters) according to manufacturer’s protocol. The eluent was lyophilized and reconstituted with 25 µL of 2% ACN, 0.1% FA in water prior to LC-MS analysis.

The insoluble tau was quantified as previously described with modifications.^38^ The samples were reconstituted in 5 µL 1.5 M tris-HCl pH 8.8 buffer, sonicated briefly and centrifuged at 10,000 *g* for 5 min at 4°C. 25 µL urea buffer (8 M urea, 5 mM TCEP, pH 8.5) was added and internal tau protein standards were added, samples were reduced and alkylated as described above. The samples were diluted to 250 µL using 50 mM TEABC to dilute the urea below 1 M and centrifuged at room temperature for 15 min at 10,000 *g*. The proteins were digested using LysC enzyme (300 ng per sample) for 4 hours at 37°C, followed by trypsin digestion (500 ng) for 18 hours at 37°C. Post enzymatic digestion, the samples were briefly centrifuged, and digestion enzymes were quenched by TFA to a final concentration of 1% (pH <2 was verified after TFA addition). The samples were spiked with AQUA peptide standards and desalted using Oasis µElution HLB plate (Waters) as described above. The eluent was lyophilized and reconstituted in 25 µL of 2% ACN, 0.1% FA in water prior to LC-MS analysis.

The soluble as well as the insoluble tau peptides were analyzed on a Vanquish Neo UHPLC (ThermoFisher Scientific) coupled to an Orbitrap Exploris 480 (ThermoFisher Scientific, San Jose, CA). The tau peptides were separated using the same LC-MS configuration as described above. Tau peptides were eluted from the column with a gradient of 4-8% of buffer B for 10 min, then 8-20% of buffer B for another 8 min before ramping up to 95% buffer B in next 2 min and cleaning the column for another 2 min. Twenty-one non-phosphorylated brain tau peptides were quantified by comparison with the corresponding isotopomer signals from the ^15^N internal standard using Skyline software (version 20.2).^44^ Four phosphorylated peptides (pT181, pT205, pS208 and pT217) were quantified. To assess the phosphorylation occupancy of specific sites in tau from soluble brain homogenates and insoluble extracts, we measured the extent of phosphorylation on each site as described previously.^40,43^ Briefly, endogenous peak area of the phosphorylated peptides were normalized with the corresponding peak area of the non-phosphorylated peptide. Absolute quantitation was performed by using heavy to light ratio of the internal stable isotope standards (AQUA peptides) for each phosphorylated and non-phosphorylated peptide.

### Statistical Analysis

Welch two sample t-tests were used to estimate statistical differences in insoluble Aβ isoforms between either the gantenerumab or solanezumab treatment arms and the DIAD observational cohort for each brain region, and *p* values were adjusted for false discovery rate (FDR) control by the Benjamini–Hochberg procedure. Baseline demographic characteristics were summarized using means and standard deviations for normally distributed continuous variables, medians with interquartile ranges for skewed continuous variables, and counts and percentages for categorical variables. Cohort diagnostic information was summarized by treatment group using the last available record for each participant. Aβ biomarker values in both soluble and insoluble fractions were summarized across brain regions and treatment groups.

Linear mixed-effects (LME) models were used to compare Aβ biomarkers across treatment groups for each brain region. The LME model included the fixed effects of treatment group (solanezumab, gantenerumab, mutation carriers, and non-carriers), brain regions (eight), their interaction, and individual-level random intercepts to account for the correlation across multiple brain regions within the same participant. If the interaction term was not statistically significant, it was removed from the model.

To evaluate the association between gantenerumab exposure and each Aβ biomarker, we employed a second linear mixed-effects model. This analysis was restricted to mutation carriers who received gantenerumab treatment and had cortical and subcortical brain regions with sufficient data availability. This model included the total cumulative gantenerumab dose as a fixed effect and individual-level random intercepts. All tests were two-sided, and P < 0.05 was considered as significant. Statistical analyses and figures generation were conducted with R software and SAS 9.4

## Results

### Gantenerumab-treated DIAD brains exhibit low insoluble Aβ burden; solanezumab-treatment has no significant impact on DIAD brain Aβ burden

Despite the growing use of ATTs,^45^ their effects on the biochemical pools of Aβ in the human brain remain incompletely understood. Distinct biochemical pools of AD brain-derived soluble and insoluble Aβ contain multiple Aβ species (Aβ_38_-Aβ_43_).^36,37,46,47^ The detergent soluble and insoluble parenchymal plaques are primarily composed of Aβ_42_, followed by Aβ_40_, and Aβ_42_ is known to highly correlate with the Aβ_mid-domain_ (the common peptide marker for amyloid aggregates/fibrils).^36,37,39–41,47^ Therefore, quantitative assessment of these Aβ proteoforms in brain tissue homogenates should reveal any evidence of ATT effects on amyloid plaque pathology. To investigate the impact of prior gantenerumab or solanezumab treatment on the biochemical pools of Aβ, we measured both detergent soluble and insoluble Aβ proteoforms in multiple brain regions from drug-treated DIAN-TU-001 trial participants, untreated mutation carriers (DIAN Obs-MC participants), and age-relevant, cognitively normal controls (NC) with modest or no AD neuropathologic change (Table 3, Table 4 and Supplementary Table 1).

**Table 3.**
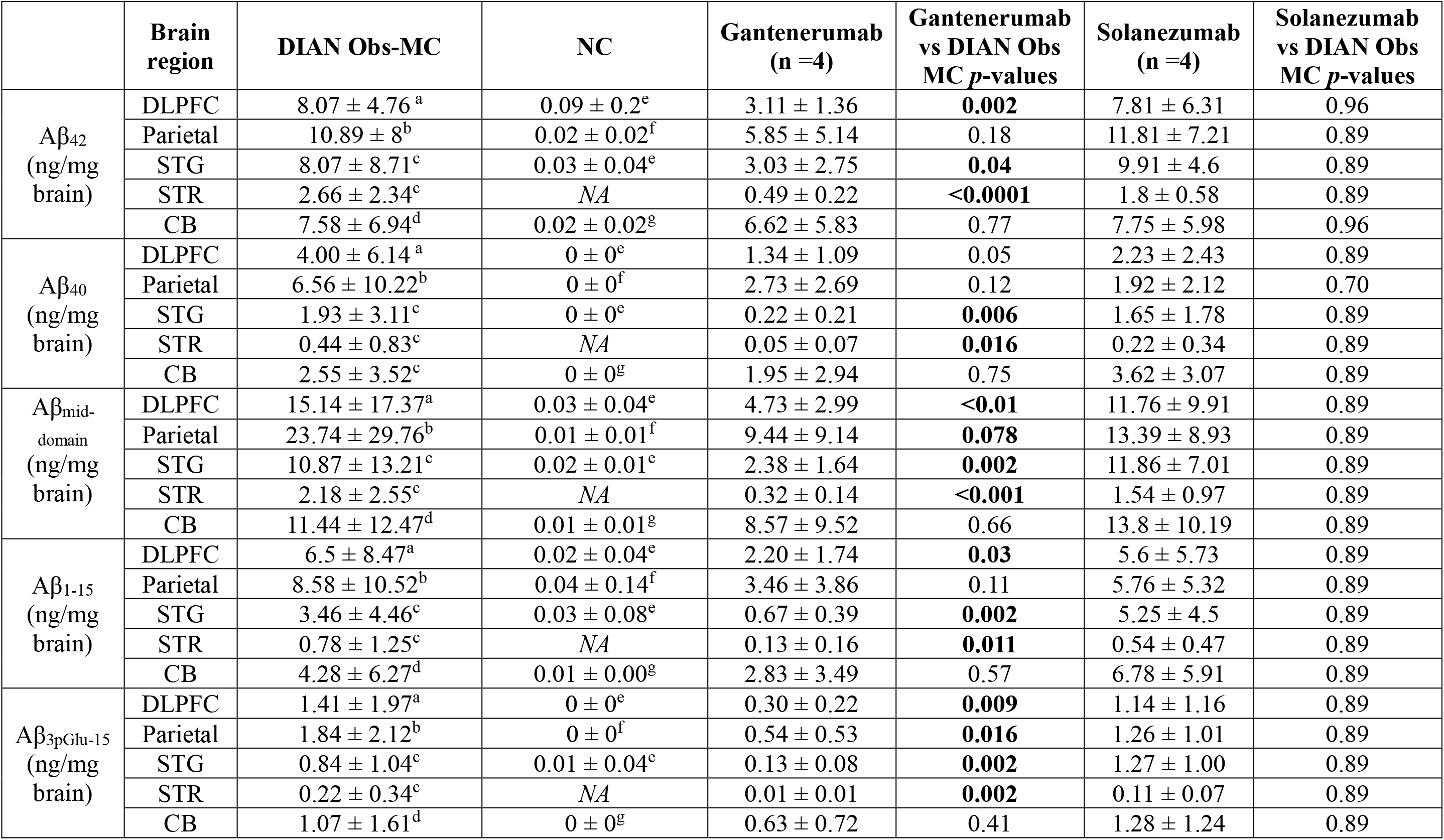
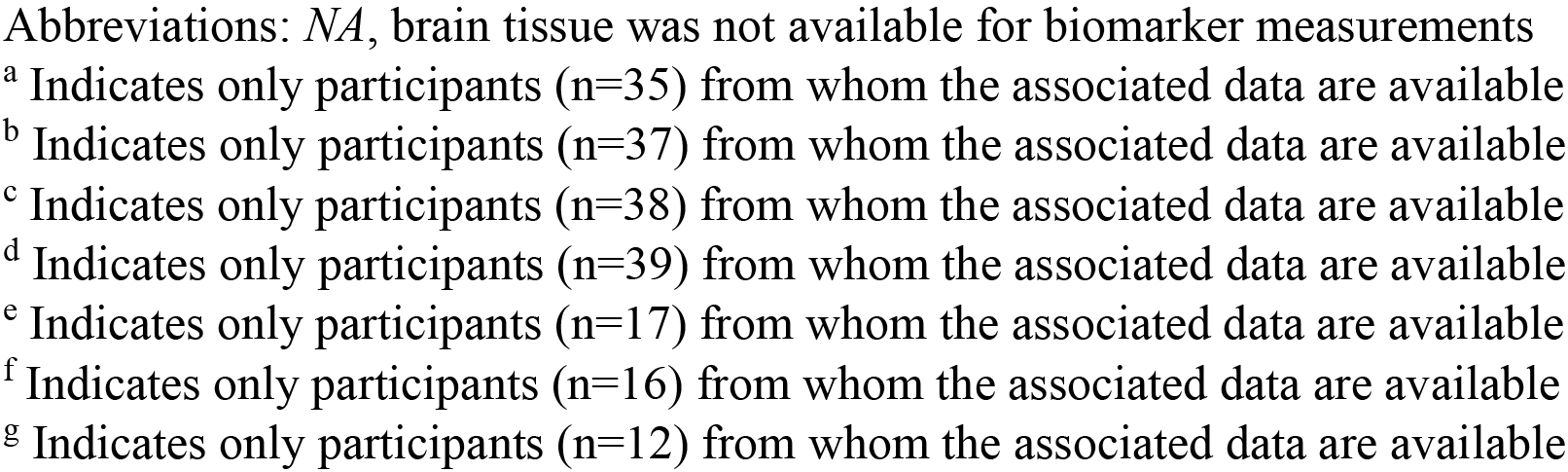
Quantitative measures of the formic acid insoluble Aβ isoforms from the different brain regions investigated from NC, DIAN Obs-MC and DIAN-TU drug treated patients. All data are represented as mean (± standard deviation). FDR adjusted *p*-values are provided for the Welch’s test for each biomarker between treatment group compared to DIAN Obs-MC.

| | Brain region | DIAN Obs-MC | NC | Gantenerumab (n =4) | Gantenerumab vs DIAN Obs MC $p$-values | Solanezumab (n =4) | Solanezumab vs DIAN Obs MC $p$-values |
| --- | --- | --- | --- | --- | --- | --- | --- |
| A$\beta_{42}$ (ng/mg brain) | DLPFC | 8.07 ± 4.76<sup>a</sup> | 0.09 ± 0.2<sup>e</sup> | 3.11 ± 1.36 | **0.002** | 7.81 ± 6.31 | 0.96 |
| | Parietal | 10.89 ± 8<sup>b</sup> | 0.02 ± 0.02<sup>f</sup> | 5.85 ± 5.14 | 0.18 | 11.81 ± 7.21 | 0.89 |
| | STG | 8.07 ± 8.71<sup>c</sup> | 0.03 ± 0.04<sup>e</sup> | 3.03 ± 2.75 | **0.04** | 9.91 ± 4.6 | 0.89 |
| | STR | 2.66 ± 2.34<sup>c</sup> | NA | 0.49 ± 0.22 | **<0.0001** | 1.8 ± 0.58 | 0.89 |
| | CB | 7.58 ± 6.94<sup>d</sup> | 0.02 ± 0.02<sup>g</sup> | 6.62 ± 5.83 | 0.77 | 7.75 ± 5.98 | 0.96 |
| A$\beta_{40}$ (ng/mg brain) | DLPFC | 4.00 ± 6.14<sup>a</sup> | 0 ± 0<sup>e</sup> | 1.34 ± 1.09 | 0.05 | 2.23 ± 2.43 | 0.89 |
| | Parietal | 6.56 ± 10.22<sup>b</sup> | 0 ± 0<sup>f</sup> | 2.73 ± 2.69 | 0.12 | 1.92 ± 2.12 | 0.70 |
| | STG | 1.93 ± 3.11<sup>c</sup> | 0 ± 0<sup>e</sup> | 0.22 ± 0.21 | **0.006** | 1.65 ± 1.78 | 0.89 |
| | STR | 0.44 ± 0.83<sup>c</sup> | NA | 0.05 ± 0.07 | **0.016** | 0.22 ± 0.34 | 0.89 |
| | CB | 2.55 ± 3.52<sup>c</sup> | 0 ± 0<sup>g</sup> | 1.95 ± 2.94 | 0.75 | 3.62 ± 3.07 | 0.89 |
| A$\beta_{\text{mid-domain}}$ (ng/mg brain) | DLPFC | 15.14 ± 17.37<sup>a</sup> | 0.03 ± 0.04<sup>e</sup> | 4.73 ± 2.99 | **<0.01** | 11.76 ± 9.91 | 0.89 |
| | Parietal | 23.74 ± 29.76<sup>b</sup> | 0.01 ± 0.01<sup>f</sup> | 9.44 ± 9.14 | **0.078** | 13.39 ± 8.93 | 0.89 |
| | STG | 10.87 ± 13.21<sup>c</sup> | 0.02 ± 0.01<sup>e</sup> | 2.38 ± 1.64 | **0.002** | 11.86 ± 7.01 | 0.89 |
| | STR | 2.18 ± 2.55<sup>c</sup> | NA | 0.32 ± 0.14 | **<0.001** | 1.54 ± 0.97 | 0.89 |
| | CB | 11.44 ± 12.47<sup>d</sup> | 0.01 ± 0.01<sup>g</sup> | 8.57 ± 9.52 | 0.66 | 13.8 ± 10.19 | 0.89 |
| A$\beta_{1-15}$ (ng/mg brain) | DLPFC | 6.5 ± 8.47<sup>a</sup> | 0.02 ± 0.04<sup>e</sup> | 2.20 ± 1.74 | **0.03** | 5.6 ± 5.73 | 0.89 |
| | Parietal | 8.58 ± 10.52<sup>b</sup> | 0.04 ± 0.14<sup>f</sup> | 3.46 ± 3.86 | 0.11 | 5.76 ± 5.32 | 0.89 |
| | STG | 3.46 ± 4.46<sup>c</sup> | 0.03 ± 0.08<sup>e</sup> | 0.67 ± 0.39 | **0.002** | 5.25 ± 4.5 | 0.89 |
| | STR | 0.78 ± 1.25<sup>c</sup> | NA | 0.13 ± 0.16 | **0.011** | 0.54 ± 0.47 | 0.89 |
| | CB | 4.28 ± 6.27<sup>d</sup> | 0.01 ± 0.00<sup>g</sup> | 2.83 ± 3.49 | 0.57 | 6.78 ± 5.91 | 0.89 |
| A$\beta_{3p\text{Glu-15}}$ (ng/mg brain) | DLPFC | 1.41 ± 1.97<sup>a</sup> | 0 ± 0<sup>e</sup> | 0.30 ± 0.22 | **0.009** | 1.14 ± 1.16 | 0.89 |
| | Parietal | 1.84 ± 2.12<sup>b</sup> | 0 ± 0<sup>f</sup> | 0.54 ± 0.53 | **0.016** | 1.26 ± 1.01 | 0.89 |
| | STG | 0.84 ± 1.04<sup>c</sup> | 0.01 ± 0.04<sup>e</sup> | 0.13 ± 0.08 | **0.002** | 1.27 ± 1.00 | 0.89 |
| | STR | 0.22 ± 0.34<sup>c</sup> | NA | 0.01 ± 0.01 | **0.002** | 0.11 ± 0.07 | 0.89 |
| | CB | 1.07 ± 1.61<sup>d</sup> | 0 ± 0<sup>g</sup> | 0.63 ± 0.72 | 0.41 | 1.28 ± 1.24 | 0.89 |

Abbreviations: *NA*, brain tissue was not available for biomarker measurements
<sup>a</sup> Indicates only participants (n=35) from whom the associated data are available
<sup>b</sup> Indicates only participants (n=37) from whom the associated data are available
<sup>c</sup> Indicates only participants (n=38) from whom the associated data are available
<sup>d</sup> Indicates only participants (n=39) from whom the associated data are available
<sup>e</sup> Indicates only participants (n=17) from whom the associated data are available
<sup>f</sup> Indicates only participants (n=16) from whom the associated data are available
<sup>g</sup> Indicates only participants (n=12) from whom the associated data are available

**Table 4.**
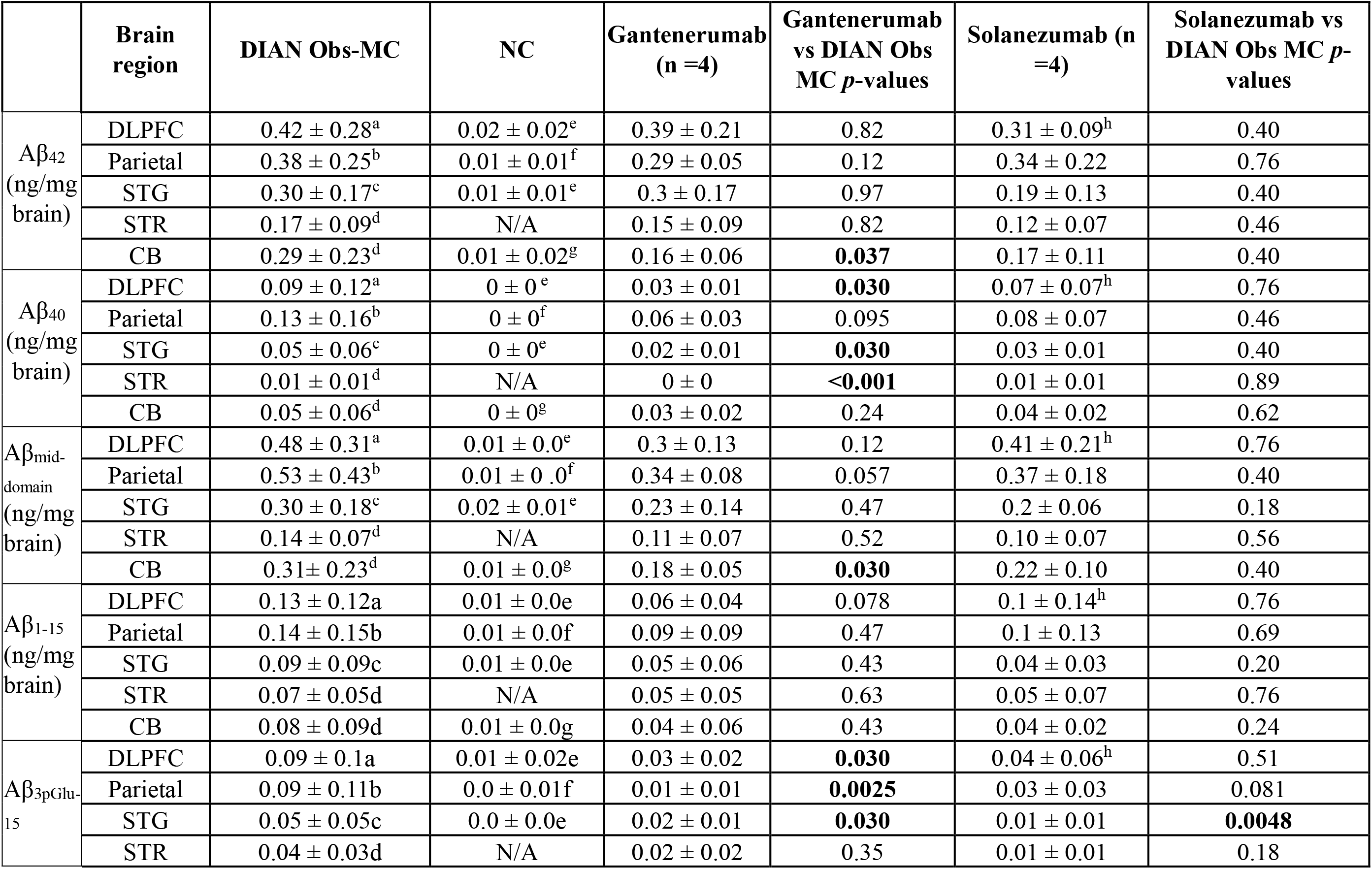
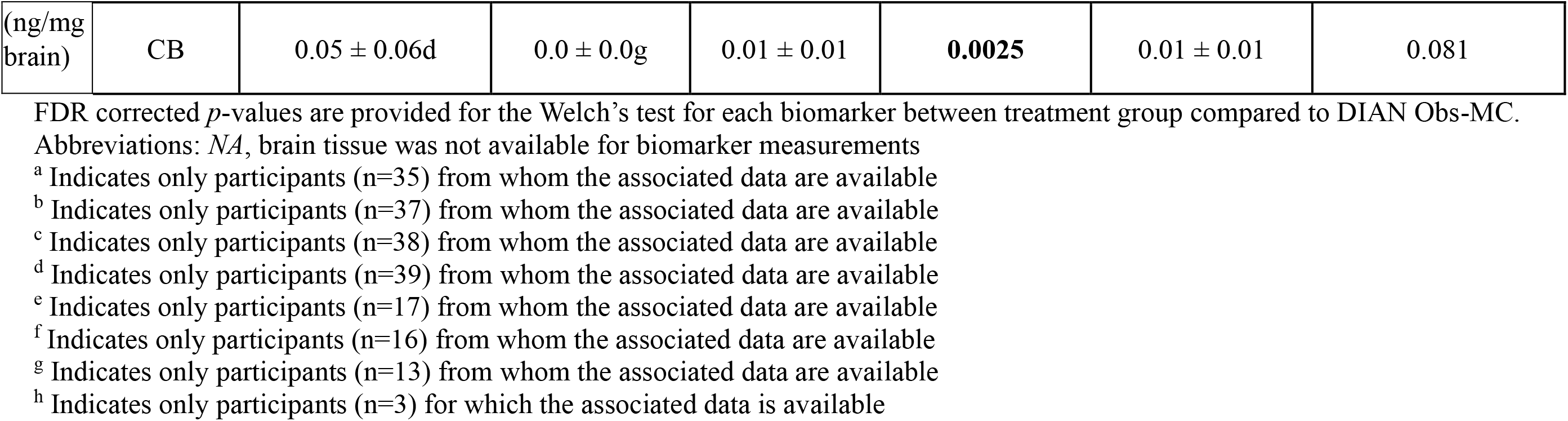
Quantitative measures of the soluble Aβ isoforms from the different brain regions investigated from NC, Obs-MC and drug treated patients. All data are represented as mean (± standard deviation). FDR adjusted *p*-values are provided for the Welch’s test for each biomarker between treatment group compared to DIAN Obs-MC.

| | Brain region | DIAN Obs-MC | NC | Gantenerumab (n =4) | Gantenerumab vs DIAN Obs MC $p$-values | Solanezumab (n =4) | Solanezumab vs DIAN Obs MC $p$-values |
| --- | --- | --- | --- | --- | --- | --- | --- |
| A$\beta_{42}$ (ng/mg brain) | DLPFC | 0.42 ± 0.28<sup>a</sup> | 0.02 ± 0.02<sup>e</sup> | 0.39 ± 0.21 | 0.82 | 0.31 ± 0.09<sup>h</sup> | 0.40 |
| | Parietal | 0.38 ± 0.25<sup>b</sup> | 0.01 ± 0.01<sup>f</sup> | 0.29 ± 0.05 | 0.12 | 0.34 ± 0.22 | 0.76 |
| | STG | 0.30 ± 0.17<sup>c</sup> | 0.01 ± 0.01<sup>e</sup> | 0.3 ± 0.17 | 0.97 | 0.19 ± 0.13 | 0.40 |
| | STR | 0.17 ± 0.09<sup>d</sup> | N/A | 0.15 ± 0.09 | 0.82 | 0.12 ± 0.07 | 0.46 |
| | CB | 0.29 ± 0.23<sup>d</sup> | 0.01 ± 0.02<sup>g</sup> | 0.16 ± 0.06 | **0.037** | 0.17 ± 0.11 | 0.40 |
| A$\beta_{40}$ (ng/mg brain) | DLPFC | 0.09 ± 0.12<sup>a</sup> | 0 ± 0<sup>e</sup> | 0.03 ± 0.01 | **0.030** | 0.07 ± 0.07<sup>h</sup> | 0.76 |
| | Parietal | 0.13 ± 0.16<sup>b</sup> | 0 ± 0<sup>f</sup> | 0.06 ± 0.03 | 0.095 | 0.08 ± 0.07 | 0.46 |
| | STG | 0.05 ± 0.06<sup>c</sup> | 0 ± 0<sup>e</sup> | 0.02 ± 0.01 | **0.030** | 0.03 ± 0.01 | 0.40 |
| | STR | 0.01 ± 0.01<sup>d</sup> | N/A | 0 ± 0 | **<0.001** | 0.01 ± 0.01 | 0.89 |
| | CB | 0.05 ± 0.06<sup>d</sup> | 0 ± 0<sup>g</sup> | 0.03 ± 0.02 | 0.24 | 0.04 ± 0.02 | 0.62 |
| A$\beta_{\text{mid-domain}}$ (ng/mg brain) | DLPFC | 0.48 ± 0.31<sup>a</sup> | 0.01 ± 0.0<sup>e</sup> | 0.3 ± 0.13 | 0.12 | 0.41 ± 0.21<sup>h</sup> | 0.76 |
| | Parietal | 0.53 ± 0.43<sup>b</sup> | 0.01 ± 0.0<sup>f</sup> | 0.34 ± 0.08 | 0.057 | 0.37 ± 0.18 | 0.40 |
| | STG | 0.30 ± 0.18<sup>c</sup> | 0.02 ± 0.01<sup>e</sup> | 0.23 ± 0.14 | 0.47 | 0.2 ± 0.06 | 0.18 |
| | STR | 0.14 ± 0.07<sup>d</sup> | N/A | 0.11 ± 0.07 | 0.52 | 0.10 ± 0.07 | 0.56 |
| | CB | 0.31 ± 0.23<sup>d</sup> | 0.01 ± 0.0<sup>g</sup> | 0.18 ± 0.05 | **0.030** | 0.22 ± 0.10 | 0.40 |
| A$\beta_{1-15}$ (ng/mg brain) | DLPFC | 0.13 ± 0.12<sup>a</sup> | 0.01 ± 0.0<sup>e</sup> | 0.06 ± 0.04 | 0.078 | 0.1 ± 0.14<sup>h</sup> | 0.76 |
| | Parietal | 0.14 ± 0.15<sup>b</sup> | 0.01 ± 0.0<sup>f</sup> | 0.09 ± 0.09 | 0.47 | 0.1 ± 0.13 | 0.69 |
| | STG | 0.09 ± 0.09<sup>c</sup> | 0.01 ± 0.0<sup>e</sup> | 0.05 ± 0.06 | 0.43 | 0.04 ± 0.03 | 0.20 |
| | STR | 0.07 ± 0.05<sup>d</sup> | N/A | 0.05 ± 0.05 | 0.63 | 0.05 ± 0.07 | 0.76 |
| | CB | 0.08 ± 0.09<sup>d</sup> | 0.01 ± 0.0<sup>g</sup> | 0.04 ± 0.06 | 0.43 | 0.04 ± 0.02 | 0.24 |
| A$\beta_{3p\text{Glu-15}}$ | DLPFC | 0.09 ± 0.1<sup>a</sup> | 0.01 ± 0.02<sup>e</sup> | 0.03 ± 0.02 | **0.030** | 0.04 ± 0.06<sup>h</sup> | 0.51 |
| | Parietal | 0.09 ± 0.11<sup>b</sup> | 0.0 ± 0.01<sup>f</sup> | 0.01 ± 0.01 | **0.0025** | 0.03 ± 0.03 | 0.081 |
| | STG | 0.05 ± 0.05<sup>c</sup> | 0.0 ± 0.0<sup>e</sup> | 0.02 ± 0.01 | **0.030** | 0.01 ± 0.01 | **0.0048** |
| | STR | 0.04 ± 0.03<sup>d</sup> | N/A | 0.02 ± 0.02 | 0.35 | 0.01 ± 0.01 | 0.18 |
| (ng/mg<br>brain) | CB | $0.05 \pm 0.06^d$ | $0.0 \pm 0.0^g$ | $0.01 \pm 0.01$ | **0.0025** | $0.01 \pm 0.01$ | 0.081 |

Abbreviations: *NA*, brain tissue was not available for biomarker measurements
<sup>a</sup> Indicates only participants (n=35) from whom the associated data are available
<sup>b</sup> Indicates only participants (n=37) from whom the associated data are available
<sup>c</sup> Indicates only participants (n=38) from whom the associated data are available
<sup>d</sup> Indicates only participants (n=39) from whom the associated data are available
<sup>e</sup> Indicates only participants (n=17) from whom the associated data are available
<sup>f</sup> Indicates only participants (n=16) from whom the associated data are available
<sup>g</sup> Indicates only participants (n=13) from whom the associated data are available
<sup>h</sup> Indicates only participants (n=3) for which the associated data is available

We first analyzed the impact of gantenerumab or solanezumab treatment on insoluble Aβ_42_ levels (Figure 2 and Supplementary Figure S2-S3). According to a mixed linear model across all brain regions, insoluble Aβ_42_ levels were significantly lower in gantenerumab treated participants compared to untreated DIAN Obs-MC participants (estimate (β): −4.29 ng/mg brain, standard error (SE): 1.98 ng/mg brain, *p* = 0.03) (Table 5). Regionally, gantenerumab treated participants showed ∼ 60% lower insoluble Aβ_42_ levels in the cortical regions (∼ 61 % in dorsolateral prefrontal cortex [DLPFC], ∼ 46 % in parietal cortex and 62 % in superior temporal gyrus [STG]) and ∼ 82% lower insoluble Aβ_42_ levels in the striatum (caudate and putamen), but no significant difference in the cerebellum, compared to untreated DIAN Obs-MC participants (Table 3, Figure 2, Supplementary Figure S4). We also evaluated the effect of total gantenerumab dose on the insoluble Aβ_42_ burden across cortical and subcortical brain regions and observed a non-significant trend towards a dose-dependent decrease (β: −0.00012, SE: 0.000073, t-value= −1.66, *p* = 0.11) (Supplementary Figure S5 and Supplementary Table 2). However, even with the highest total dose of gantenerumab (48420 mg) the insoluble Aβ_42_ levels (1.99, 1.08, 0.82, 0.37 ng/mg brain in DLPFC, parietal cortex, STG and striatum, respectively) did not reach levels comparable to those of the DIAD non-carrier brains (range 0.02-0.09 ng/mg brain Aβ_42_ across cortical brain regions [no striatal tissue samples were available for the NC]) (Figure 2 and Table 3). Samples from solanezumab treated participants showed no statistically significant difference in insoluble Aβ_42_ levels from DIAN Obs-MC samples in a mixed linear model (β: 0.23 ng/mg brain, SE: 1.98 ng/mg brain, *p* = 0.91) (Table 5), nor any significant differences for any individual brain region tested (Figure 2, Table 3 and Supplementary Figure S4)

**Table 5.** Results of the mixed model analysis for the postmortem cohort investigating the group changes of the respective biomarkers in different brain regions for each drug.

| Analyte | Estimate | SE | DF | t-value | p value |
| --- | --- | --- | --- | --- | --- |
| Insoluble Aβ<sub>42</sub> | | | | | |
| Gantenerumab vs DIAN Obs-MC | -4.299 | 1.983 | 239 | -2.17 | **0.03** |
| Solanezumab vs DIAN Obs-MC | 0.233 | 1.988 | 239 | 0.12 | 0.91 |
| Insoluble Aβ<sub>40</sub> | | | | | |
| Gantenerumab vs DIAN Obs-MC | -1.950 | 1.615 | 239 | -1.21 | 0.23 |
| Solanezumab vs DIAN Obs-MC | -0.914 | 1.619 | 239 | -0.56 | 0.57 |
| Insoluble Aβ<sub>mid-domain</sub> | | | | | |
| Gantenerumab vs DIAN Obs-MC | -8.283 | 4.863 | 239 | -1.70 | 0.09 |
| Solanezumab vs DIAN Obs-MC | -1.697 | 4.876 | 239 | -0.35 | 0.72 |
| Insoluble Aβ<sub>1-15</sub> | | | | | |
| Gantenerumab vs DIAN Obs-MC | -3.060 | 2.007 | 239 | -1.52 | 0.13 |
| Solanezumab vs DIAN Obs-MC | 0.333 | 2.013 | 239 | 0.17 | 0.87 |
| Insoluble Aβ<sub>3pGlu-15</sub> | | | | | |
| Gantenerumab vs DIAN Obs-MC | -8.283 | 4.863 | 239 | -1.7 | 0.09 |
| Solanezumab vs DIAN Obs-MC | -1.697 | 4.876 | 239 | -0.35 | 0.73 |
| Soluble Aβ<sub>42</sub> | | | | | |
| Gantenerumab vs DIAN Obs-MC | -0.055 | 0.068 | 240 | -0.80 | 0.42 |
| Solanezumab vs DIAN Obs-MC | -0.078 | 0.069 | 240 | -1.13 | 0.26 |
| Soluble Aβ<sub>40</sub> | | | | | |
| Gantenerumab vs DIAN Obs-MC | -0.035 | 0.028 | 240 | -1.23 | 0.22 |
| Solanezumab vs DIAN Obs-MC | -0.019 | 0.028 | 240 | -0.67 | 0.50 |
| Soluble Aβ<sub>mid-domain</sub> | | | | | |
| Gantenerumab vs DIAN Obs-MC | -0.119 | 0.074 | 240 | -1.62 | 0.11 |
| Solanezumab vs DIAN Obs-MC | -0.089 | 0.074 | 240 | -1.20 | 0.23 |
| Soluble Aβ<sub>1-15</sub> | | | | | |
| Gantenerumab vs DIAN Obs-MC | -0.043 | 0.028 | 240 | -1.51 | 0.13 |
| Solanezumab vs DIAN Obs-MC | -0.033 | 0.029 | 240 | -1.14 | 0.26 |
| Soluble Aβ<sub>3pGlu-15</sub> | | | | | |
| Gantenerumab vs DIAN Obs-MC | -0.043 | 0.021 | 240 | -2.07 | **0.04** |
| Solanezumab vs DIAN Obs-MC | -0.039 | 0.021 | 240 | -1.83 | 0.07 |

Abbreviations: DF, degrees of freedom; SE, standard error

Next, we examined levels of insoluble Aβ_40_ and Aβ_16-27_ (Aβ_mid-domain_); according to the mixed linear model, measured across all brain areas, these showed a decreasing trend with gantenerumab treatment compared to untreated DIAN Obs-MC, albeit, without reaching statistical significance (Aβ_40_, β: −1.95 ng/mg brain, SE: 1.61 ng/mg brain, *p* = 0.23; and Aβ_16-27_, β: −8.28 ng/mg brain, SE: 4.73 ng/mg brain, *p* = 0.09) (Table 5). Regionally, gantenerumab-treated participants exhibited ∼ 66 % lower levels of insoluble Aβ_40_ in DLPFC and ∼ 89 % lower insoluble Aβ_40_ levels in STG and striatum, but no differences in the parietal cortex and cerebellum, compared to untreated DIAN Obs-MC participants (Table 3, Figure 2, and Supplementary Figure S4A); they also showed lower insoluble Aβ_mid-domain_ levels in DLPFC (∼69%), STG (∼78%), and striatum (∼ 85 %), but no significant differences in the parietal cortex or cerebellum (Table 3, Figure 2, and Supplementary Figure S4A). These results align with those for insoluble Aβ_42,_ providing additional evidence that gantenerumab treatment reduces total insoluble amyloid plaque burden. In contrast, insoluble levels of these Aβ_40_ and Aβ_mid-domain_ species were not lower in any brain regions of solanezumab-treated participants relative to untreated DIAN Obs-MC participants, corroborating that solanezumab does not clear insoluble fibrillar amyloid plaques (Supplementary Figure S4C).

Next, we investigated levels of soluble Aβ proteoforms Aβ_40,_ and Aβ_42,_ and the Aβ_mid-domain_ (Figure 3 and Table 4). Gantenerumab treated participants exhibited significantly lower levels of soluble Aβ_40_ in the DLPFC (∼67%), STG (∼60%), and striatum (∼ 100 %) when compared to untreated DIAN Obs-MC participants (Figure 3, Table 4 and Supplementary Figure S4B), but not in the parietal cortex or cerebellum. Gantenerumab treated participants also showed significantly lower levels of soluble Aβ_42_ and Aβ_mid-domain_ in the cerebellum (∼45% and ∼42%, respectively) but not in other brain areas. No significant solanezumab-associated difference was detected in the level of any of these three soluble Aβ proteoforms in any brain region (Figure 3, Tables 4 and 5, and Supplementary Figure S4D).

**Figure 3.**
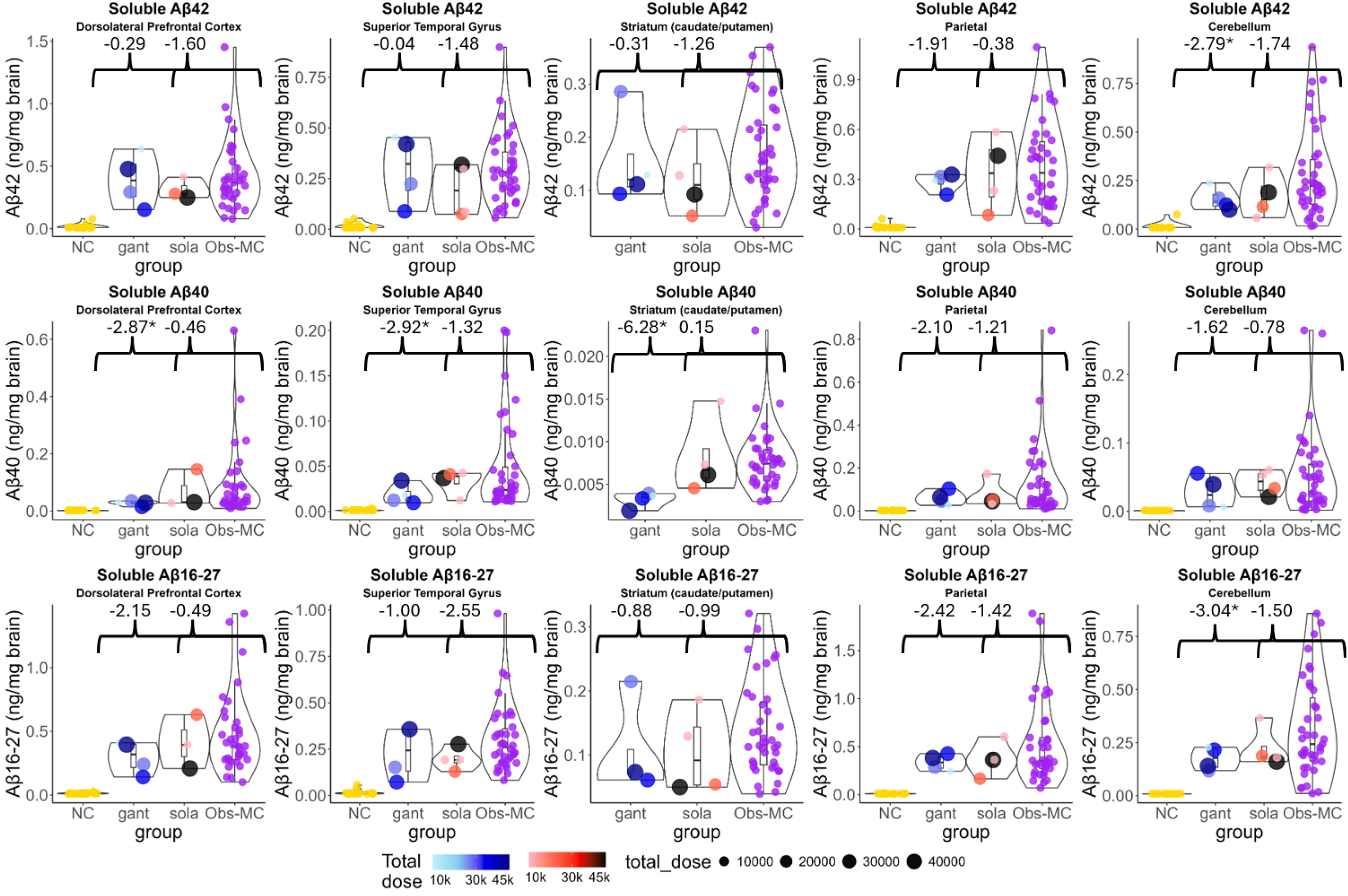
Soluble Aβ_40_ isoform levels in the brains of symptomatic DIAD mutation carriers are lowered with gantenerumab treatment. Scatter plots of quantitative measures of the soluble (top) Aβ_x42_ (ng/mg brain), (middle) Aβ_40_ (ng/mg brain) and (bottom) Aβ_mid-domain_ (ng/mg brain), in different brain regions (dorsolateral prefrontal cortex, superior temporal gyrus, striatum, parietal cortex, and cerebellum) of postmortem brains of age-relevant, cognitively normal non-DIAD controls (NC, n =18), DIAD mutation carriers treated with gantenerumab (gant, n=4) or solanezumab (sola, n=4), and untreated observational DIAD mutation carriers (DIAN Obs-MC, n=40). The total cumulative dose (mg) of the drug received in the clinical trial for gantenerumab (blue) and solanezumab (red) is highlighted in color gradient for each participant. Asterisks denote FDR adjusted *p*-values < 0.05 (*) associated with Welch two-sample t-tests comparing Aβ isoforms between the Obs-MC group and the gantenerumab arm or the solanezumab arm.

### Insoluble and soluble Aβ N-terminus post-translational modifications are altered in gantenerumab-treated brains

Truncations and post-translational modifications (PTMs) of the Aβ N-terminus resulting from distinct amyloid processing pathways play an important role in amyloid aggregation/fibril formation.^47–49^ Therefore, we measured the unmodified N-terminus of Aβ (Aβ_1-15_) and the pyroglutamate modified N-terminus of Aβ (Aβ_3pGlu-15_) (Figure 2A) to investigate the potential effects of gantenerumab and solanezumab on these biochemically distinct Aβ pools (Figure 4). Regionally, we found lower insoluble Aβ_1-15_ levels in gantenerumab-treated participant DLPFC (∼66%), STG (∼80%) and striatum (∼80%) compared to untreated DIAN Obs-MC participants, and no differences in parietal cortex or cerebellum (Table 3, Figure 4, Supplementary Figure S4A). For insoluble Aβ_3pGlu-15_, we found lower levels in gantenerumab-treated participants across neocortical brain regions (∼ 79 % in DLPFC, ∼ 85% in STG and ∼ 71% in parietal cortex) and the striatum (∼ 95 %), and no difference in cerebellum, compared to the untreated DIAN-Obs MC participant brains (Table 3 and Supplementary Figure S4A). According to the mixed linear model, soluble Aβ_3pGlu-15_ levels overall were significantly lower in gantenerumab-treated participants (β: −0.04 ng/mg brain, SE: 0.02 ng/mg brain, *p* = 0.04) compared to untreated DIAN Obs-MC participants (Table 5); regionally, these lower levels of soluble Aβ_3pGlu-15_ were detected in DLPFC (∼67%), parietal cortex (∼89%), STG (∼60%), and cerebellum (∼80%) (Table 4 and Supplementary Figure S4B). This finding suggests that amyloid clearance by gantenerumab can affect both insoluble and soluble levels of this post-translationally modified Aβ proteoform.

**Figure 4.**
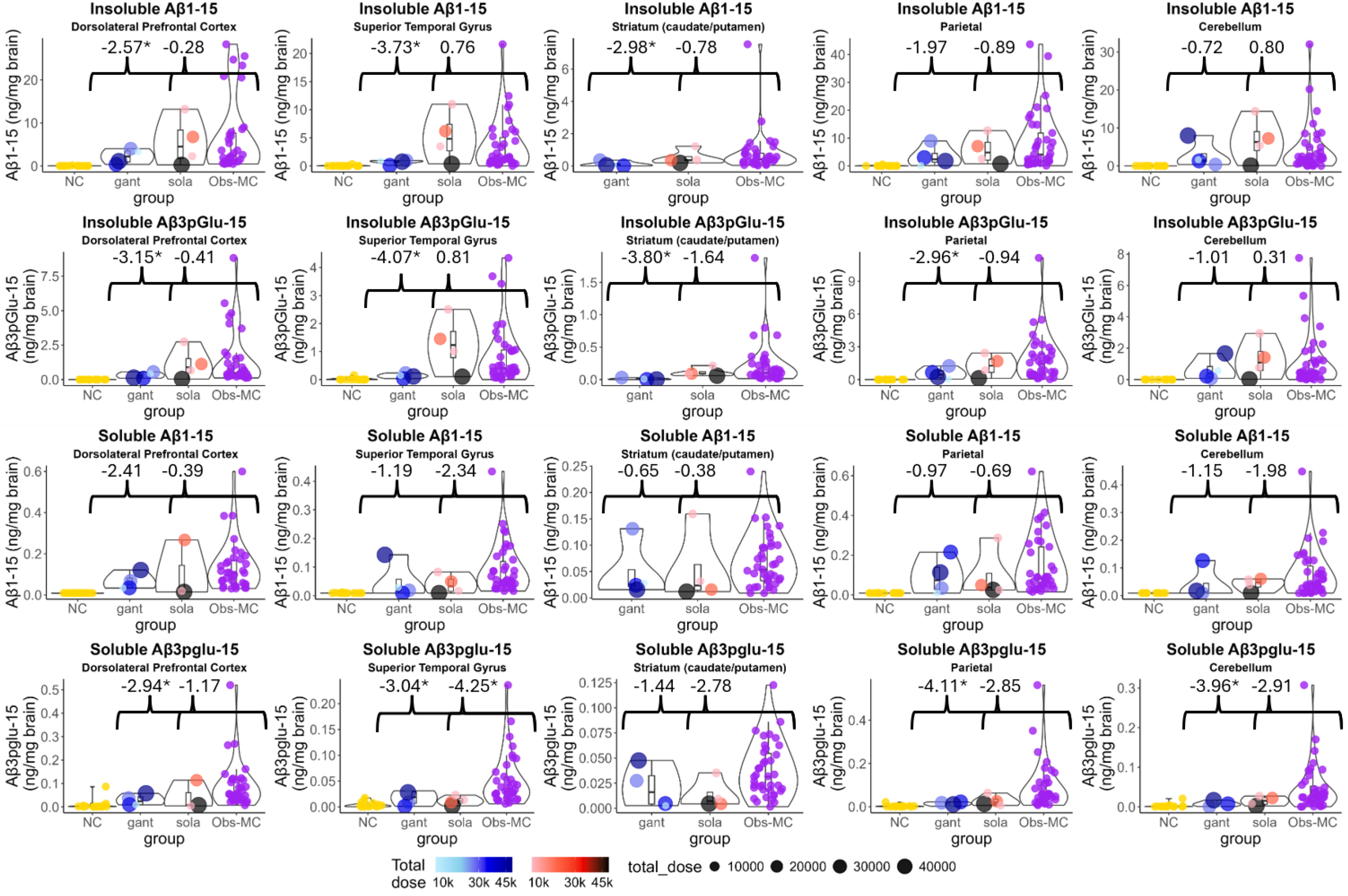
Reduction of insoluble and soluble Aβ N-terminus brain levels in symptomatic DIAD mutation carriers treated with gantenerumab. Scatter plots of quantitative measures of the insoluble (top row) Aβ_1-x_ (ng/mg brain), (second row) pGlu3-Aβ (ng/mg brain), (third row), soluble Aβ_1-x_ (ng/mg brain) and (bottom row) soluble pGlu3-Aβ (ng/mg brain), in different brain regions (dorsolateral prefrontal cortex, superior temporal gyrus, striatum, parietal cortex, and cerebellum) from the postmortem brains of age-relevant, cognitively normal non-DIAD controls (NC, n =18), DIAD mutation carriers treated with gantenerumab (gant, n=4) or solanezumab (sola, n=4), and untreated observational DIAD mutation carriers (DIAN Obs-MC, n=40). The total cumulative dose (mg) of the drug received in the clinical trial for gantenerumab (blue) and solanezumab (red) is highlighted in color gradient for each participant. Asterisks denote FDR adjusted *p*-values < 0.05 (*) associated with Welch two-sample t-tests comparing Aβ isoforms between the Obs-MC group and the gantenerumab arm or the solanezumab arm.

In contrast, though solanezumab-treated participants did show a lower level of soluble Aβ_3pGlu-15_ in the STG (∼80%) compared to untreated DIAN-Obs MC participants (Supplementary Figure S4D), they showed no differences in insoluble or soluble Aβ_1-15_ and Aβ_3pGlu-15_ levels in the mixed linear models (Table 5) nor in any other individual brain region (Table 3, Table 4, Figure 4, Supplementary Figure S4C).

### Neurofibrillary tangles burden remain unchanged, gantenerumab-treatment reduces insoluble tau phosphorylation

Recent antemortem imaging and biofluid biomarker studies have shown that gantenerumab treated participants demonstrated treatment-associated reductions in Aβ-PET tracer retention and in measures of CSF Aβ42/40 and phosphorylated tau (pT153, pT181, pT217).^29,30,50^ We also investigated whether these ATTs may have impacted NFTs, formed by fibrillar hyperphosphorylated tau protein, derived from the brain tissue homogenates. We and others have demonstrated that sarkosyl insoluble fractions derived from AD brain samples with NFTs contain tau species hyperphosphorylated at many sites (including pT181, pT205, pS208 and pT217) as well as N-terminal and C-terminal truncated, microtubule binding region (MTBR) rich tau species;^38,51–54^ the more abundant of these insoluble MTBR fragments span residues 299-317, 306-317 and 354-369 (Supplementary Figure S6). The measured absolute levels of pathological insoluble tau (MTBR-tau354-369) across brain regions in DIAN Obs-MC participants, presented as average values in ng per wet weight (ng/mg) of brain tissue, showed that cortical regions have higher levels (∼ 330, DLPFC; ∼ 300, parietal cortex and ∼ 250, STG) relative to the striatum (with 10-fold lower levels [∼ 20]), and that the cerebellum has far lower levels (< 1) (Figure 5); these findings are wholly consistent with immunohistochemical assessments of DIAN Obs-MC brain donations using antibodies raised against hyperphosphorylated tau epitopes (e.g., PHF-1) and used to stage and/or quantify burdens of tauopathy in AD.^32,55^

**Figure 5.**
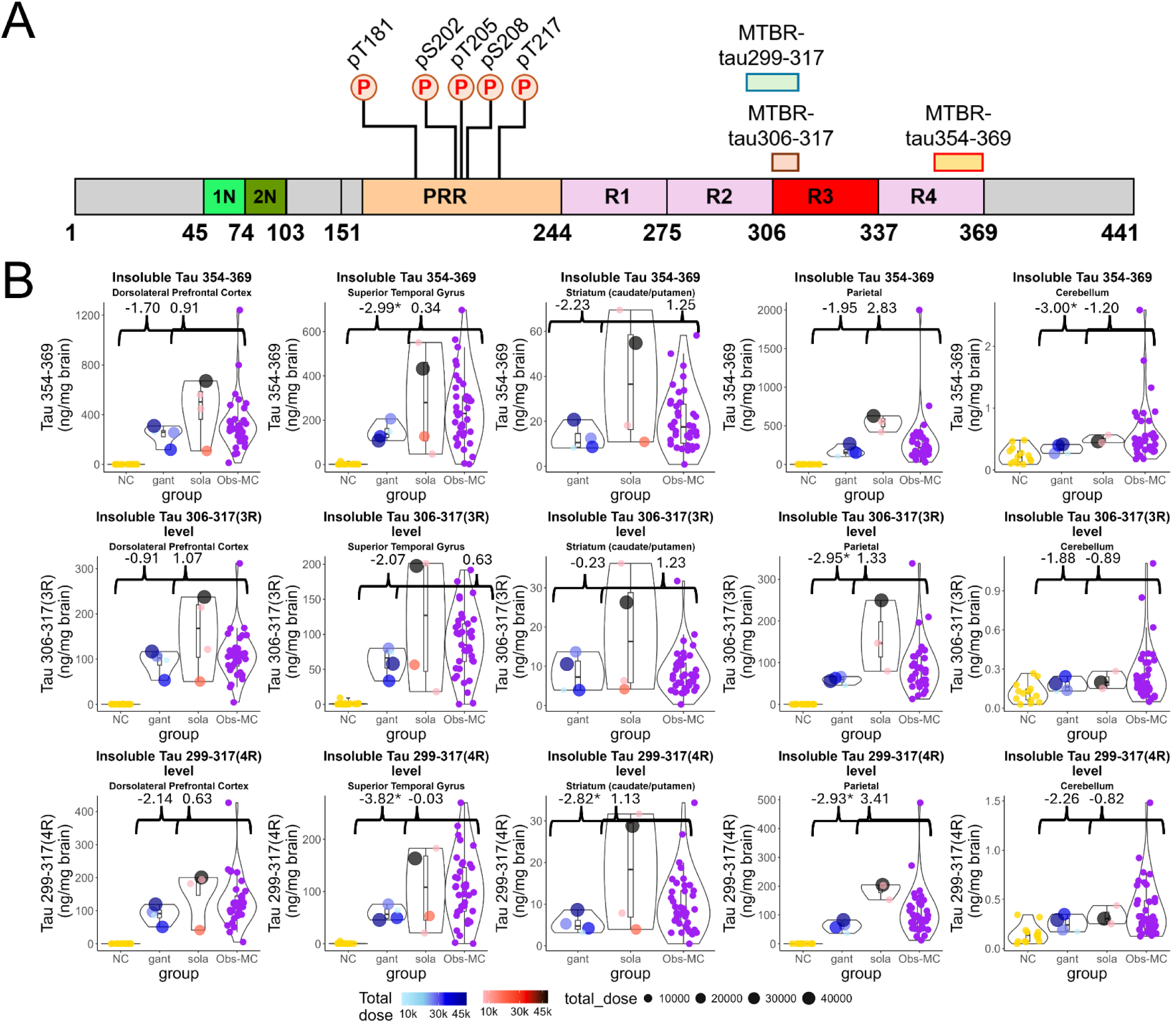
Insoluble MTBR-tau in the brains of symptomatic DIAD mutation carriers are not altered after anti-amyloid drug treatment. (A) Schematic representation of canonical 2N4R tau (441 amino acids) sequence with the N-terminal inserts, proline rich region (PRR) and the four microtubule binding repeat regions (MTBR) highlighted. Three tryptic MTBR-tau peptides, MTBR-tau299-317, MTBR-tau306-317 and MTBR-tau354-369 are depicted as green, red and orange bars, respectively, and five ptau sites (pT181, pS202, pT205, pS208 and pT217) are highlighted. (B) Scatter plots of quantitative measures of the sarkosyl insoluble MTBR-tau354-369 (top), MTBR-tau306-317 (3R) (middle) and MTBR-tau299-317 (4R) (bottom) levels (ng/mg brain) in different brain regions (dorsolateral prefrontal cortex, superior temporal gyrus, striatum, parietal cortex, and cerebellum) of postmortem brains of age-relevant, cognitively normal non-DIAD controls (NC, n=18), DIAD mutation carriers treated with gantenerumab (gant, n=4) or solanezumab (sola, n=4), and untreated observational DIAD mutation carriers (DIAN Obs-MC, n=40). The total cumulative dose (mg) of the drug received in the clinical trial for gantenerumab (blue) and solanezumab (red) is highlighted in color gradient for each participant. Asterisks denote FDR adjusted *p*-values < 0.05 (*) associated with Welch two-sample t-tests comparing Aβ isoforms between the Obs-MC group and the gantenerumab arm or the solanezumab arm.

We then compared the levels of these insoluble tau proteoforms across brain regions from gantenerumab-treated, solanezumab-treated, and untreated DIAN Obs-MC participant brain donors. With mixed linear modeling, we found no differences in insoluble levels of MTBR-tau354-369 between gantenerumab-treated participants and untreated DIAN-Obs MC participants (Supplementary Table 3). Regionally, however, we found lower levels of insoluble MTBR-tau354-369 in the STG and cerebellum, and lower levels of insoluble MTBR-tau299-317 (4R tau) in the parietal cortex, STG and striatum of gantenerumab treated participants, compared to untreated DIAN Obs-MC participants (Table 6, Figure 5, Supplementary Figure S7A). We found no regional differences in the insoluble phosphorylated tau-T217 (pT217) levels in gantenerumab other than in striatum (∼ 50 % lower) and solanezumab treated participants compared to those from the untreated DIAN Obs-MC participants (Supplementary Figure S7-8 and Supplementary Table 4). Additionally, relative to untreated DIAN Obs-MC participants, gantenerumab-treated participants exhibited ∼ 43 to 67% lower insoluble pT205 levels in the DLPFC, parietal cortex, STG, and striatum, ∼ 58 to 67% lower insoluble pS208 levels in the DLPFC, parietal cortex, and STG, and ∼ 51 to 71% lower insoluble pT181 levels in the DLPFC, parietal cortex, STG, striatum, and cerebellum (Figure 6, Table 6 and Supplementary Figure S7A). Taken together, these findings suggest that some tau phosphorylations (e.g., pT181, pT205 and pT208) may be reduced by gantenerumab treatment in parallel with amyloid plaque clearance, whereas established NFTs (represented by insoluble MTBR-tau species) may be relatively more stable and less-affected by gantenerumab treatment.

**Figure 6.**
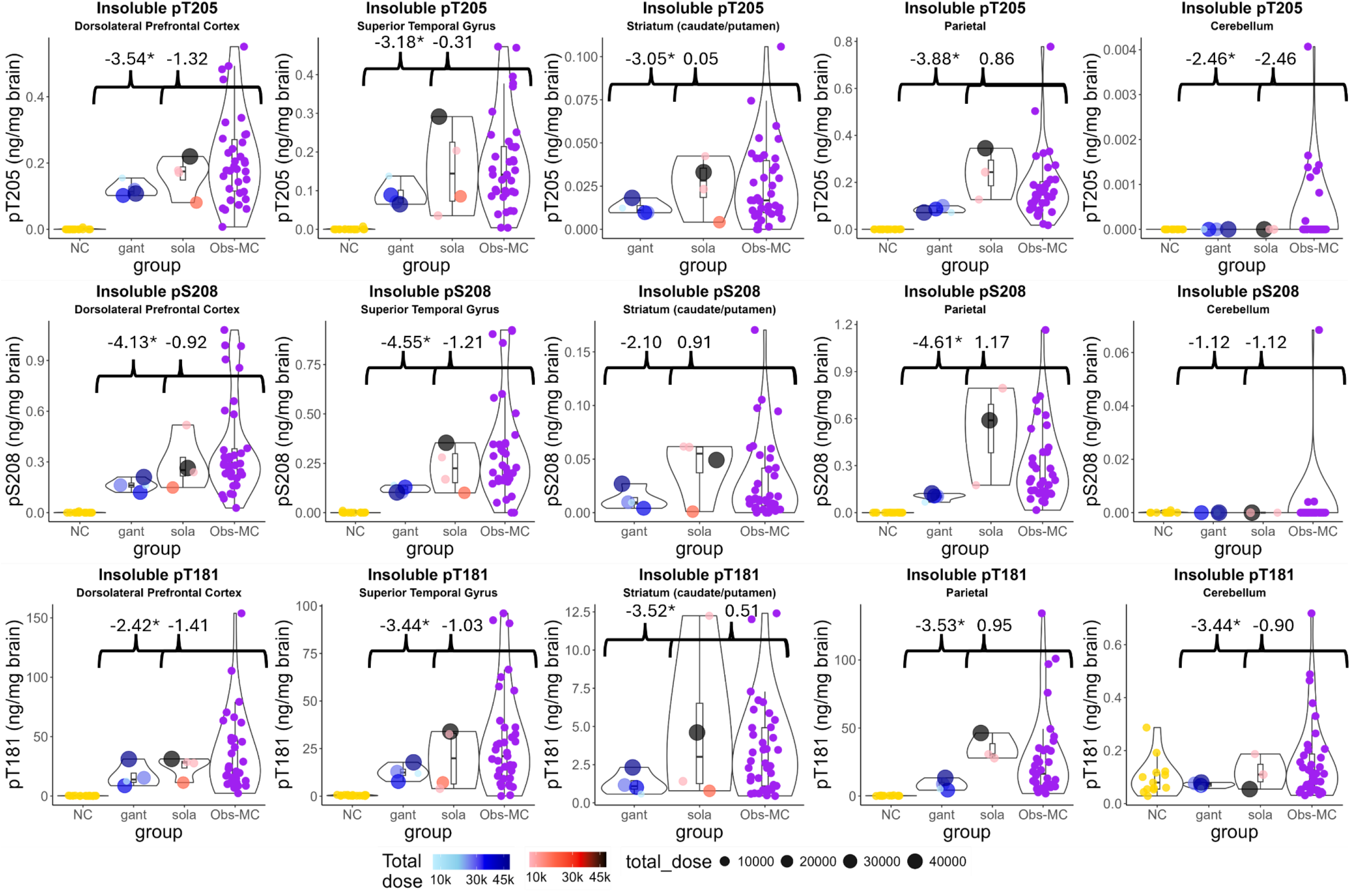
Insoluble phosphorylated tau levels in the brains of symptomatic DIAD mutation carriers show regional alteration with gantenerumab treatment. Scatter plots of quantitative measures of the sarkosyl insoluble pT205 (top row), pS208 (middle row) and pT181 (4R) (bottom row) levels (ng/mg brain) in different brain regions (dorsolateral prefrontal cortex, superior temporal gyrus, striatum, parietal cortex, and cerebellum) of postmortem brains of age-relevant, cognitively normal non-DIAD controls (NC, n=18), DIAD mutation carriers treated with gantenerumab (gant, n=4) or solanezumab (sola, n=4), and untreated observational DIAD mutation carriers (DIAN Obs-MC, n=40). The total cumulative dose (mg) of the drug received in the clinical trial for gantenerumab (blue) and solanezumab (red) is highlighted in color gradient for each participant. Asterisks denote FDR adjusted *p*-values < 0.05 (*) associated with Welch two-sample t-tests comparing Aβ isoforms between the Obs-MC group and the gantenerumab arm or the solanezumab arm.

**Table 6.**
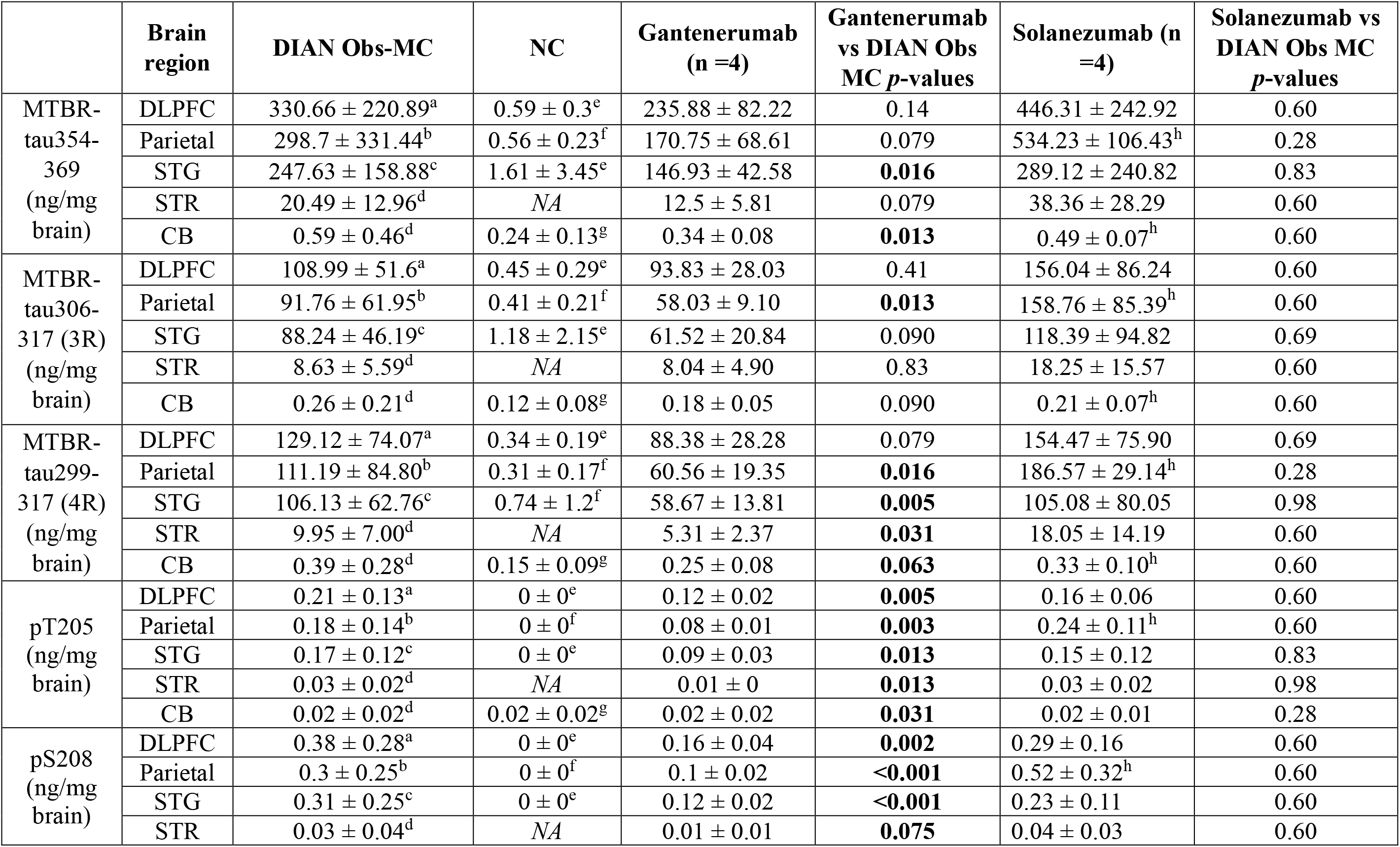
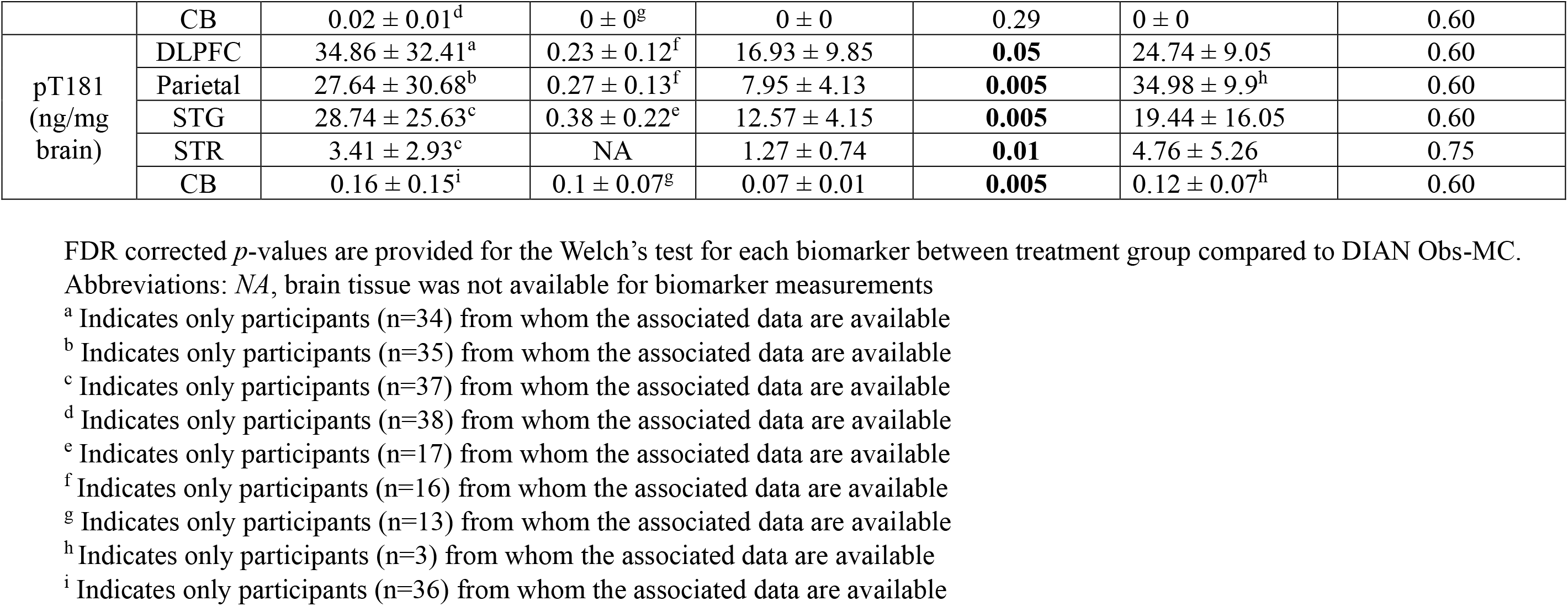
Quantitative measures of the sarkosyl insoluble unphosphorylated (MTBR) and phosphorylated tau isoforms from the different brain regions investigated from NC, DIAN Obs-MC and DIAN-TU drug treated patients. All data are represented as mean (± standard deviation). FDR adjusted *p*-values are provided for the Welch’s test for each biomarker between treatment group compared to DIAN Obs-MC.

Whereas CSF total tau (independent of phosphorylation status), pT205 and MTBR-tau243 increase close to clinical symptom onset and correlate with tau-PET positivity,^43,50,56,57^ recent biomarker studies suggest that increased CSF pT217, pT231 and pT181 reflect amyloid-plaque pathology in the brain, as they correlate with but change even earlier in the disease course than Aβ-PET positivity in both DIAD and LOAD.^17,58^ We thus investigated the soluble phospho-tau (pT181, pS202, pT205, pS208 and pT217) and MTBR species across the same brain regions (Supplementary Figure S8 and Supplementary Table 4).

No significant differences were found in the soluble levels of any of the five phospho-tau species nor of any of the three MTBR species among gantenerumab-treated participants, solanezumab-treated participants, and untreated DIAD participants, in any brain regions (Supplementary Figure S8 and Supplementary Table 4).

## Discussion

In this study, we investigated how Aβ and tau proteoforms in the brains of DIAD participants were affected by treatment with gantenerumab or solanezumab, two Aβ-targeting monoclonal antibodies, in the DIAN-TU-001 trial and gantenerumab open label extension.^15^ During that trial, Aβ PET and CSF biomarkers were used to monitor target engagement and downstream treatment responses.^15,29,59^ Retrospective postmortem studies have examined the impact of mild to extensive amyloid plaque removal in clinical trials that involved either active (AN1792) or passive immunization (aducanumab, lecanemab, gantenerumab, solanezumab) of AD patients,^31,33–35,60,61^ but have generally relied on immunohistochemical plaque or tangle-area-fraction measures. A biochemically resolved comparison of insoluble and soluble Aβ isoforms along with tau pathology across multiple brain regions in symptomatic AD patients treated for a long duration with ATTs is limited. Our study is unique as it examined the impact of these drugs on numerous specific Aβ and tau protein species in symptomatic DIAD participants, for whom the predicted ages of symptom onset – and the actual ages of onset – were known. This allowed for direct comparisons to a large number of untreated symptomatic DIAD mutation carriers (DIAN Obs study participants) and age-relevant, cognitively normal, non-carrier controls with minimal or no AD neuropathologic change.

Previously, a quantitative immunohistochemical evaluation of these gantenerumab-treated and solanezumab-treated DIAN-TU-001 participant brain donors (in comparison to untreated DIAN Obs affiliated brain donors) provided direct evidence of dose-dependent removal of Aβ deposits by gantenerumab, without striking alterations in tauopathy, microgliosis, or astrocytosis.^32^ In this companion study of those treated participants, we utilized biochemical fractionation and high resolution mass spectrometry (LC-MS) assays to understand the impact of these drug treatments on the abundance and aggregation states and post-translational modifications of numerous Aβ and tau protein species that are known to change in AD brain.^36–38,41^ We observed that gantenerumab significantly reduced the amounts of insoluble Aβ proteoforms (Aβ_42_, Aβ_40_ and Aβ_mid-domain_) in striatum and neocortical regions. Activation of microglial phagocytic states around parenchymal amyloid plaques has been reported in both actively and passively immunized brains, correlating with antibody response and Aβ removal.^35^ As gantenerumab is known to bind to insoluble Aβ plaques and aggregates and trigger microglial phagocytosis of the amyloid plaques,^26^ our finding of decreased insoluble Aβ peptides is not unexpected.

The incomplete and variable reduction of insoluble Aβ peptides observed among the gantenerumab-treated participants in this study is also unsurprising, as Aβ PET and CSF biomarker measurements during the DIAN-TU-001 trial were consistent with lowered but persistently abnormal amyloid plaque burdens, and the dosages received during the DIAN-TU-001 trial and subsequent gantenerumab open label extension period differed for each participant.^26^ Recent reports on neuropathological examinations of anti-amyloid targeting therapies (aducanumab) also indicate spatial and regional variability in amyloid removal.^31,33,34^ While amyloid reduction with gantenerumab is noted, the total amyloid burden does not reach the levels observed in postmortem brains from age-relevant non-DIAD cognitively normal controls with low or no ADNC. Amyloid levels on Aβ-PET in GRADUATE I and GRADUATE II clinical trials, which evaluated gantenerumab in the context of late-onset sporadic AD, have also demonstrated similar ∼ 50 % reduction at week 116 compared to the placebo groups.^62^ The apparent preferential gantenerumab-associated removal of insoluble Aβ peptides from the striatum relative to cortical regions was also anticipated, as similar findings have been reported by others in response to bapineuzumab,^63^ and we reported preferential striatal plaque clearance in response to gantenerumab in our prior immunohistochemical (IHC) analyses of these treated participants.^32^ This regional variability and uneven reduction of Aβ burden by gantenerumab as measured by Aβ-PET, with prominent reduction in subcortical regions, has also been demonstrated in DIAN-TU-001 participants.^30^ This preferential clearance from the striatum could be modulated by enhanced blood brain barrier permeability of the drug and subsequent clearance from the brain,^64,65^ or morphological/biochemical differences between striatal versus cortical amyloid plaques, but the exact mechanism remains uncertain and needs further investigation.

One unexpected finding was no apparent difference in the levels of any insoluble Aβ peptide isoforms (Aβ_42_, Aβ_40_, _Aβ1-15_, Aβ_3pGlu-15_ and Aβ_mid-domain_) in the cerebelli of gantenerumab-treated participants compared to those of untreated DIAD participants, despite our previous finding of lower gantenerumab-associated Aβ IHC area fractions in the cerebellar gray matter. ^26^ Although these potentially conflicting results seem difficult to reconcile, we suggest that, in DIAD, as in sporadic AD, postmortem cerebellar amyloid burden (both vascular and parenchymal) can be highly variable regionally within a cerebellum; they can also differ across cases with identical ADNC stages, and even among family members who share a DIAD mutation. In our previous IHC study, ^26^ cerebellar samples from gantenerumab-treated participants uniformly represented dorsomedial cerebellar cortex from the fixed left hemibrain; in this study, cerebellar samples were derived from more lateral cerebellar regions of the frozen right hemibrain. These sampling differences alone may account for the different results. Another potential contributing factor is the retention / greater representation of leptomeningeal vessels and CAA in our cerebellar gray matter preparations in comparison to our cortical gray matter preparations; it is possible that vascular deposits of Aβ peptide and parenchymal deposits of Aβ peptide contribute differently to IHC area fraction than they do to biochemical / mass spectrometric measures of insoluble Aβ species. Reconciling these findings with greater certainty will require further evaluation in the future.^66^

In contrast to gantenerumab-treated participants, solanezumab-treated participants did not show lower levels of insoluble Aβ peptide species in any brain regions, in comparison to untreated DIAD participants. This difference may reflect the fundamentally different mechanism of action of the two drugs. Unlike gantenerumab, solanezumab is intended to reduce Aβ deposition in the brain by binding and removing soluble monomeric Aβ. Upon examination of soluble Aβ levels, we found significantly smaller amounts of soluble Aβ_40_ and post-translationally modified Aβ_3pglu_ in the striatum and neocortical regions of gantenerumab-treated participants, with lower amounts of soluble Aβ_42_ and Aβ_mid-domain_ limited to the cerebellum. Consistent with our brain findings, the CSF Aβ_42/40_ ratio was noted to increase in response to gantenerumab during the DIAN-TU-001 trial.^60,67^ CSF Aβ_40_ levels were also decreased in the GRADUATE I and GRADUATE II phase 3 studies,^68^ a change interpreted to represent a compensatory increase in soluble Aβ due to a shift of the equilibrium between soluble and insoluble Aβ pools. Aβ_40_ is the dominant amyloid proteoform depositing in cerebral amyloid angiopathy (CAA)^41^ and vascular pathology is thought to play a role in amyloid-related imaging abnormalities (ARIA),^69,70^ a vascular side effect associated with ATTs. Whether the robust regional reductions of insoluble and soluble Aβ_40_ levels in gantenerumab treated patients could influence vascular pathology, CAA, and ARIA needs further investigation. It is particularly noteworthy that soluble levels of Aβ_3pglu_, a species formed by enzymatic post-translational modification of Aβ, were lower in association with gantenerumab treatment, as accumulated/deposited Aβ, rather than new cleavage from APP, likely determines steady state levels of soluble Aβ_3pglu_ in the brain. It is also worth noting that Aβ_3pglu_ concentrations in the brains of gantenerumab-treated participants do not appear to have regressed and risen towards the levels observed in untreated DIAN Obs MC participants, even after treatment-to-expiration gaps of two years.

One key question from the limited neuropathological studies related to anti-amyloid targeting clinical trials has been whether there is substantial or partial regional reduction of tau tangle pathology after moderate to extensive removal of plaques.^31,71,72^ These postmortem studies have reported varying degree of ‘clearing’ of tau immunoreactivity and lower tau-containing dystrophic neurites in amyloid plaque free regions.^34,60,67^ However, all these patients had progressed to advanced Braak stage (V/VI), suggesting the spread of tau pathology continued despite persistent amyloid plaque removal.^60^ Recent neuropathological investigations on aducanumab treated brains show that brain regions with extensive parenchymal plaque clearance are associated with decreased neuritic phospho-tau, but NFT (as defined by PHF-1 and AT8 staining and tangle staging) remain unaltered.^33,34^ We thus evaluated how the DIAN-TU-001 amyloid-targeting therapies influence tauopathy that is arguably ‘downstream’ of Aβ plaque deposition. In our study, gantenerumab-treated participants showed fairly widespread lower levels of certain insoluble phosphorylated tau species (pT181, pT205 and pT208), and less widespread lower regional levels of insoluble MTBR-tau species, but no differences in insoluble levels of pT217, when compared to untreated DIAN Obs-MC participants. The significantly lower levels we observed in insoluble pT181, pT205 and pS208 levels could be consistent with partial reduction in hyperphosphorylated tau burden following sustained amyloid-plaque clearance. Since our biochemical assay of insoluble tau fraction cannot distinguish tangle-derived from neuritic-derived tau, high resolution spatial techniques will be required to investigate and resolve any compartment-specific changes. In participants who received solanezumab, there was no significant difference in any of the brain regions for these tau species except for higher levels of some MTBR species in the parietal and dorsolateral prefrontal cortices, which would be consistent with worsening tau pathology, not a reduction of tau pathology. These results suggest that, even though NFT pathology was not substantively reduced, partial lowering of amyloid plaque burden with gantenerumab had some downstream effects on insoluble levels of phosphorylated tau in these representative DIAN-TU-001 study brain donors.

Notably, the lack of change in soluble brain p-tau stands in contrast to the fluid biomarker data from the DIAN-TU trial, where gantenerumab significantly reduced CSF soluble pT181 and pT217 levels, while solanezumab had no such effect.^50^ This discrepancy suggests that brain soluble p-tau measures do not necessarily reflect CSF soluble p-tau. Whereas CSF p-tau is likely derived from a pool of tau species secreted into the brain’s extracellular interstitial fluid, soluble brain p-tau likely represents a mixture of interstitial fluid and intracellular sources.^17,73^ These two pools of p-tau may respond differently to amyloid plaque removal by gantenerumab and, in brain tissue with substantial tauopathy, intracellular p-tau levels are likely far more abundant than extracellular levels.

While our study provides valuable findings, there are limitations. It is important to consider the difference in dosages given to the participants, the duration of treatment, and the gap period of being off treatment before death caused by end-stage AD dementia and brain donation. Of these initial 4 cases from DIAN-TU-001 in each drug arm, these participants were the first to donate because they were more advanced at the time of entry into the trial, less likely to complete full treatment compared to other participants who are still actively being treated in DIAN-TU Amyloid Removal Trial, and received lower doses compared to other participants who are still active in the trial, some of whom are still asymptomatic. The first major limitation of this study is the small sample size, as it included only four participants who had been treated with each drug. Nevertheless, as we demonstrated previously,^15,32^ the DIAN-TU-001 Aβ PET and CSF biomarker data for these eight participants resemble those of their DIAN-TU-001 cohorts of origin, suggesting that they adequately represent at least these key features of the larger groups in DIAN-TU-001. Further, despite this small size, this study found significant reductions in brain tissue-derived insoluble and soluble Aβ proteoform levels that align with the corresponding CSF and Aβ PET biomarker-based findings.

Another limitation related to the small number of brain tissue specimens currently available from DIAN-TU-001 trial is the lack of a genuine DIAN-TU-001 placebo group to which to compare the drug-treated participant brain donors. In lieu of a placebo control group for this study, DIAN Obs-MC participant and DIAN-Obs family member brain donors were selected for inclusion. It seems unlikely that this alternative DIAD control group would alter the results from this study, but we acknowledge that a true placebo group would be preferred.

A third limitation stems from the interval between last drug treatment and neuropathologic evaluation – greater than one year for all but one of the drug-treated participants, during which substantial re-accumulation of amyloid pathology and continued advancement of non-amyloid AD pathologic features may have occurred. However, this continued progression would have countered the reductions in insoluble and soluble Aβ proteoform levels that were observed; therefore, this limitation actually bolsters the key finding of this study.

A fourth limitation, inherent to all such neuropathologic studies, is the bias of excluding the living (reverse survivorship bias). As reported recently, a subset of gantenerumab-treated DIAN-TU-001 participants who were cognitively normal at baseline, continued gantenerumab treatment in the open label extension period for an average of 8 years, and received the largest cumulative doses, have experienced significant reductions in their anticipated rate of cognitive decline, with approximate 50 % reduction in the risk of onset and progression of clinical dementia.^14^ We cannot exclude the possibility that gantenerumab-treated DIAN-TU-001 participants who received larger doses and remain vital have experienced even more favorable changes in their AD neuropathologic status (including tauopathy burden) than those included in this study, who died due to complications associated with “end-stage” dementia and with severe ADNC.

In summary, this is the first comprehensive study using high resolution mass spectrometry that details the differences in Aβ and tau burden in different brain regions of DIAD participants treated with either gantenerumab or solanezumab, in comparison to untreated DIAD mutation-carriers and non-DIAD normal controls. This study provides quantitative evidence for the biochemical changes induced by these drugs in symptomatic DIAD patients. Our results demonstrate that the anti-amyloid targeting monoclonal antibody gantenerumab can alter AD pathology by decreasing insoluble and soluble levels of Aβ proteoforms, and even insoluble levels of some forms of phosphorylated tau, from neocortical and striatal brain regions. With the advent of United States Food and Drug Administration (FDA) approved anti-amyloid targeting therapies, patients with biomarker evidence of brain amyloid and mild cognitive impairment or mild AD dementia, and without other exclusionary features, are being treated with these drugs (currently, lecanemab and donanemab) in clinics across the US.^45,74,75^ In the future, to complement neuroimaging and fluid biomarker studies of these patients before, during, and after treatment, neuropathologic examination, supplemented by the methods used in this study, may provide a deeper understanding of the neuropathological changes associated with current amyloid-targeting therapies. Such insights may guide improvements to these AD treatment strategies so that, one day, we might arrest, or even prevent, AD.

## Data Availability

Data access to the DIAN–TU trial data will follow the policies of the DIAN–TU data access policy, which complies with the guidelines established by the Collaboration for Alzheimer’s Prevention. Patient-related data not included in the paper were generated as part of a clinical trial and may be subject to patient confidentiality. Any data and materials that can be shared will be released via a data/material sharing agreement. Requests to access the DIAN–TU-001 trial data can be made at https://dian.wustl.edu/our-research/for-investigators/diantu-investigator-resources/. Data are available by request to the DIAN–Obs (https://dian.wustl.edu/our-research/observational-study/dian-observational-study-investigator-resources/data-request-terms-and-instructions/).

## Acknowledgements

We gratefully acknowledge the outstanding commitment of the participants, family members, and caregivers whose participation was critical to the success of the DIAN-Obs and DIAN-TU trial. We thank the DIAN-Obs, DIAN Expanded Registry and DIAN-TU research and support staff for their exceptional dedication and amazing accomplishments which ensured the success of the trial (see [DIAN Study Team](https://dian.wustl.edu/about/team/)). Research reported in this publication was supported by the National Institute on Aging of the National Institutes of Health under Award Numbers U01AG042791, U01AG042791-S1 (FNIH and Accelerating Medicines Partnership), R1AG046179, R01AG053267-S1. This study was also supported by the Goizueta ADRC at Emory University (National Institute of Aging of the National Institutes of Health under Award Number P30 AG066511). The content is solely the responsibility of the authors and does not necessarily represent the official views of the National Institutes of Health. The research for the DIAN-TU-001 trial, solanezumab and gantenerumab drug arms, was also supported by the Alzheimer’s Association, Eli Lilly and Company, F. Hoffman LaRoche Ltd., Avid Radiopharmaceuticals (a wholly owned subsidiary of Eli Lilly and Company), GHR Foundation, an anonymous organization, Cogstate, and Signant. The DIAN-TU has received funding from the DIAN-TU Pharma Consortium. This manuscript has been reviewed by DIAN-TU Study investigators for scientific content and consistency of data interpretation with previous DIAN-TU Study publications. Data collection and sharing for this project were supported by The Dominantly Inherited Alzheimer Network (DIAN, U19AG032438) funded by the National Institute on Aging (NIA), the Alzheimer’s Association (SG-20-690363-DIAN), the German Center for Neurodegenerative Diseases (DZNE), the Raul Carrea Institute for Neurological Research (FLENI), the Research and Development Grants for Dementia from Japan Agency for Medical Research and Development (AMED), the Korea Dementia Research Project (HU21C0066) through the Korea Dementia Research Center, funded by the Ministry of Health and Welfare and Ministry of Science and ICT, Republic of Korea, the Spanish Institute of Health Carlos III (ISCIII), the Canadian Institutes of Health Research (CIHR), the Canadian Consortium of Neurodegeneration and Aging, the Brain Canada Foundation, and Fonds de Recherche du Québec – Santé. This manuscript has been reviewed by DIAN Study investigators for scientific content and consistency of data interpretation with previous DIAN Study publications. We acknowledge the altruism of the participants and their families and contributions of the DIAN research and support staff at each of the participating sites for their contributions to this study: Randall J. Bateman, Alisha J. Daniels, Laura Courtney, Eric McDade, Jorge J. Llibre-Guerra, Charlene Supnet-Bell, Chengie Xiong, Xiong Xu, Ruijin Lu, Guoqiao Wang, Yan Li, Emily Gremminger, Richard J. Perrin, Erin E. Franklin, Laura Ibanez, Gina Jerome, Elizabeth Herries, Jennifer Stauber, Bryce Baker, Matthew Minton, Carlos Cruchaga, Alison M. Goate, Alan E. Renton, Danielle M. Picarello, Tammie Benzinger, Brian A. Gordon, Russell Hornbeck, Jason Hassenstab, Jennifer Smith, Sarah Stout, Andrew J. Aschenbrenner, Celeste M. Karch, Jacob Marsh, John C. Morris, David M. Holtzman, Nicolas Barthelemy, Jinbin Xu, James M. Noble, Sarah B. Berman, Snezana Ikonomovic, Neelesh K. Nadkarni, Gregory S. Day, Neill R. Graff-Radford, Martin Farlow, Jasmeer P. Chhatwal, Takeshi Ikeuchi, Kensaku Kasuga, Yoshiki Niimi, Edward D. Huey, Stephen Salloway, Peter R. Schofield, William S. Brooks, Jacob A. Bechara, Ralph Martins, Nick C. Fox, David M. Cash, Natalie S. Ryan, Mathias Jucker, Christoph Laske, Anna Hofmann, Elke Kuder-Buletta, Susanne Graber-Sultan, Ulrike Obermueller, Johannes Levin, Yvonne Roedenbeck, Jonathan Vӧglein, Jae-Hong Lee, Jee Hoon Roh, Raquel Sanchez-Valle, Pedro Rosa-Neto, Ricardo F. Allegri, Patricio Chrem Mendez, Ezequiel Surace, Silvia Vazquez, Francisco Lopera, Yudy Milena Leon, Laura Ramirez, David Aguillon, Allan I. Levey, Erik C. B. Johnson, Nicholas T. Seyfried, John Ringman, Anne M. Fagan, Hiroshi Mori.

## Contributors

S.M., N.R.B., Y.L., E.E.F., C.M.K, B.A.G., E.M., T.L.S.B., R.J.P., and R.J.B. conceived the concept and designed the study. S.M., N.R.B., Y.L., D.Y., E.E.F., J.X., Y.H., R.C., H.F., N.S.R., C. S-B., B.A.G., E.M., T.L.S.B., C.M.K., R.J.P., and R.J.B. were responsible for acquiring, analyzing, or interpreting data. S.M., T.A., C.M.K, B.A.G., R.J.P., and R.J.B drafted the manuscript. S.M., Y.L. and D.Y. performed all the statistical analysis. All authors reviewed the manuscript critically for important intellectual content.

## Data Sharing

Data access to the DIAN–TU trial data will follow the policies of the DIAN–TU data access policy,^76^ which complies with the guidelines established by the Collaboration for Alzheimer’s Prevention. Patient-related data not included in the paper were generated as part of a clinical trial and may be subject to patient confidentiality. Any data and materials that can be shared will be released via a data/material sharing agreement. Requests to access the DIAN–TU-001 trial data can be made at https://dian.wustl.edu/our-research/for-investigators/diantu-investigator-resources/. Data are available by request to the DIAN-Obs (https://dian.wustl.edu/our-research/observational-study/dian-observational-study-investigator-resources/data-request-terms-and-instructions/).

