## Supplementary Figures for "Gantenerumab reduces brain amyloid-β proteoforms: Biochemical evaluation in dominantly inherited Alzheimer disease"

**Supplemental Information**

**Biochemical Investigations of Brain Amyloid-β and Tau in Response to Gantenerumab or Solanezumab in Dominantly Inherited Alzheimer’s Disease**

Soumya Mukherjee,^1,2^ Nicolas R. Barthélemy,^1,2^ Taraneh Atri, Yan Li,^1^  Daniel Yen,^1^ Yingxin He,^1,2^  Reid Coyle,^1^ Vitaliy Ovod,^1,2^  Erin E. Franklin,^1,3,4^ Chengjie Xiong,^1^ Guoqiao Wang,^1^ Jinbin Xu,^1^ Huifangjie Farsad,^1^ Catherine Mummery,^5^ Colin L. Masters,^6^ Raquel Sanchez-Valle,^7^ Mario Masellis,^8^ Ging-Yuek Robin Hsiung,^9,10,11^ Serge Guthier,^12^ James J. Lah,^13^ Sarah Berman,^14^ Erik D. Roberson,^15^ Ghulam Surti,^16^ Lawrence S. Honig,^17^ Roger Clarnette,^18^ Paolo Vitali,^10^ John Ringman,^19^ Thomas D. Bird,^20,21,22,23^ William S. Brooks,^24^ Nick C. Fox,^5,25^ Kazushi Suzuki,^26^ Sandra E. Black,^8,27,28,29,30^ Johannes Levin,^31,32,33^ Martin R. Farlow,^34^ Jasmeer Chhatwal,^35^ Neelum T. Aggarwal,^36^ Mathias Jucker,^37,38^ Matthew P. Frosch,^39^ Julia K. Kofler,^14^ Charles White III,^40^ C. Dirk Keene,^41^ Jie Chen,^42^ Jonathan Glass,^13^ Marla Gearing,^13^ Natalie S. Ryan,^5^ Charlene Supnet-Bell,^1^ Brian A. Gordon,^1^ Tammie L.S. Benzinger,^1^ John Sims,^43^ Roy Yaari,^43^ Tobias Bittner,^44^ Gregory Klein,^45^ Paul Delmar,^45^ John C Morris,^1^ Jorge Llibre-Guerra,^1^ David B. Clifford,^1^ Celeste M. Karch,^1,^ Eric McDade,^1^ Richard J. Perrin,^1,3,4^ Randall J. Bateman^1,2^* for the Dominantly Inherited Alzheimer Network Observational Study and Dominantly Inherited Alzheimer Network-Trials Unit^†^

1. Washington University School of Medicine, St. Louis, MO
2. The Tracy Family Stable Isotope Labeling Quantitation (SILQ) Center, Washington University in St. Louis, St. Louis, MO, USA.
3. Departments of Pathology and Immunology, Washington University School of Medicine, St. Louis, MO, USA
4. Hope Center for Neurological Disorders, Washington University, St. Louis, MO, USA;
5. Dementia Research Centre, Department of Neurodegenerative Disease, UCL Queen Square Institute of Neurology, University College London, London, UK
6. Alzheimer's Research Australia, University of Western Australia, Perth, Australia.
7. Alzheimer's Disease and Other Cognitive Disorders Unit, Neurology Service, Hospital Clínic, Institut d'Investigacións Biomèdiques August Pi I Sunyer, University of Barcelona, Barcelona, Spain.
8. Hurvitz Brain Sciences Research Program, Sunnybrook Research Institute, Toronto, Canada.
9. Division of Neurology, Departments of Medicine and Medical Sciences, University of Toronto, Toronto, Canada
10. Division of Neurology, University of British Columbia, Vancouver, BC, Canada.
11. McGill University Research Centre for Studies in Aging, McGill University, 6825 Boulevard LaSalle, Verdun, Québec, H4H 1R3, Canada.
12. Montreal Neurological Institute, McGill University, Montréal, QC, H3A 2B4, Canada.
13. Department of Neurology and Neurosurgery, McGill University, Montréal, QC, H3A 1A1, Canada.
14. Emory University, Atlanta, Georgia, USA.
15. Department of Neurology, University of Pittsburgh, Pittsburgh, PA, USA.
16. University of Alabama at Birmingham Alzheimer's Disease Research Center Birmingham Alabama USA.
17. Department of Psychiatry and Human Behavior, Warren Alpert Medical School of Brown University, Providence, RI, USA.
18. Taub Institute and Department of Neurology, Columbia University Irving Medical Center, New York, New York, USA.
19. Taub Institute, Sergievsky Center, and Department of Neurology, Columbia University Irving Medical Center, New York, NY, USA.
20. Alzheimer's Disease Research Center, Department of Neurology, Keck School of Medicine, University of Southern California, Los Angeles, California, USA.
21. Geriatrics Research Education and Clinical Center, Veterans Affairs Puget Sound Health Care System, 1660 South Columbian Way, Seattle, WA, 98108, USA.
22. Division of Gerontology and Geriatric Medicine, Department of Medicine, University of Washington, 325 9th Ave, Seattle, WA, 98104, USA.
23. Division of Medical Genetics, Department of Medicine, University of Washington, 1705 NE Pacific St, Seattle, WA, 98195, USA.
24. Department of Neurology, University of Washington Medical Center, 1959 NE Pacific Street, Seattle, WA, 98195, USA.
25. Neuroscience Research Australia, Randwick, 2031 NSW, Australia.
26. UK Dementia Research Institute at UCL, University College London, London, United Kingdom.
27. Division of Neurology, Internal Medicine, National Defense Medical College, Saitama, Japan.
28. Division of Neurology, Department of Medicine, Sunnybrook Health Sciences Centre, Toronto, ON, Canada
29. L.C. Campbell Cognitive Neurology Research Unit, Sunnybrook Health Sciences Centre, Toronto, ON, Canada
30. Department of Neurology, Ludwig-Maximilians Universität München, Munich 80539, Germany
31. German Center for Neurodegenerative Diseases (DZNE), 81377, Munich, Germany
32. Munich Cluster for Systems Neurology (SyNergy), Munich, Germany
33. Department of Neurology, Indiana University School of Medicine, Indianapolis.
34. Department of Neurology, Massachusetts General Hospital, Boston, MA, 02114, USA.
35. Rush University Medical Center Chicago Illinois USA.
36. German Center for Neurodegenerative Diseases (DZNE), Tübingen, Germany
37. Hertie-Institute for Clinical Brain Research, University of Tübingen, Tübingen, Germany.
38. Department of Neurology, Alzheimer's Disease Research Unit, Massachusetts General Hospital, Charlestown, MA 02129, USA.
39. University of Texas Southwestern, Dallas, TX, USA
40. Department of Pathology, University of California, San Diego, La Jolla, California, USA.
41. Department of Pathology and Laboratory Medicine, Emory University School of Medicine, Atlanta, GA 30322
42. Group of Neuroscience of Antioquia, GNA, Medical School, University of Antioquia, Medellin, Colombia
43. Eli Lilly and Company, Indianapolis, Indiana, USA.
44. Genentech, Inc., South San Francisco, CA, USA; F. Hoffmann-La Roche Ltd, Basel, Switzerland.
45. Pharma Research and Early Development, F Hoffmann-La Roche Ltd., Basel, Switzerland.

^†^Data used in the preparation of this article were obtained from the Dominantly Inherited Alzheimer Network (DIAN) and Dominantly Inherited Alzheimer Network Trials Unit (DIAN-TU). As such, the study team members within the DIAN and DIAN-TU contributed to the design and implementation of DIAN-TU and/or provided data but may not have participated in the analysis or writing of this report. A complete listing of the DIAN and DIAN-TU Study Team Members can be found at [dian.wustl.edu](https://dian.wustl.edu/about/team/).

**Supplementary Figure S1**. Scatter plots of quantitative estimation of the insoluble Aβ levels (ng/mg brain) in different brain regions of postmortem brains (dorsolateral prefrontal cortex, superior temporal gyrus, striatum, parietal and cerebellum) of non-DIAD normal controls (NC), DIAD mutation carriers treated with gantenerumab (gant), solanezumab (sola) and untreated DIAD observational mutation carriers (Obs-MC). The DIAD mutation status is highlighted in color for each group (*APP*, red; *PSEN1*, violet and *PSEN2*, blue). The total cumulative dose of the drug received (Table 2) in the clinical trial for gantenerumab (gant) and solanezumab (sola)-treated participants are reflected by the size of the circle.

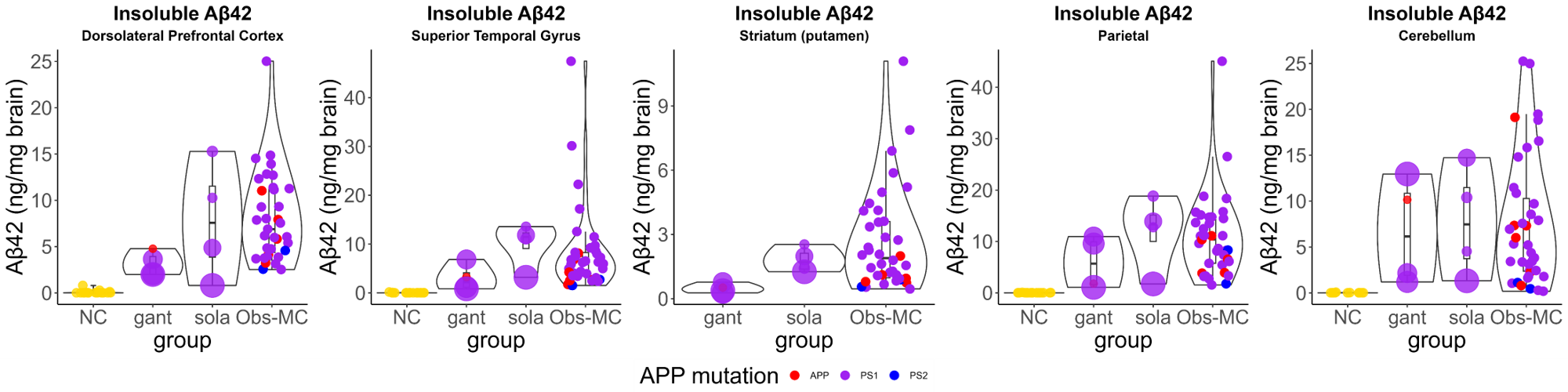

**Supplementary Figure S2.** Scatter plots of quantitative measures of the (A) insoluble and (B) soluble Aβ_42_ (ng/mg brain), (C) insoluble and (D) soluble Aβ_40_ (ng/mg brain), and (E) insoluble and (F) soluble Aβ_mid-domain_ (ng/mg brain) in all the different brain regions (dorsolateral prefrontal cortex, superior temporal gyrus, striatum, parietal cortex, cerebellum, occipital cortex, pons and thalamus) from the postmortem brains of age-relevant, cognitively normal non-DIAD controls, DIAD mutation carriers treated with gantenerumab or solanezumab and untreated observational DIAD mutation carriers .

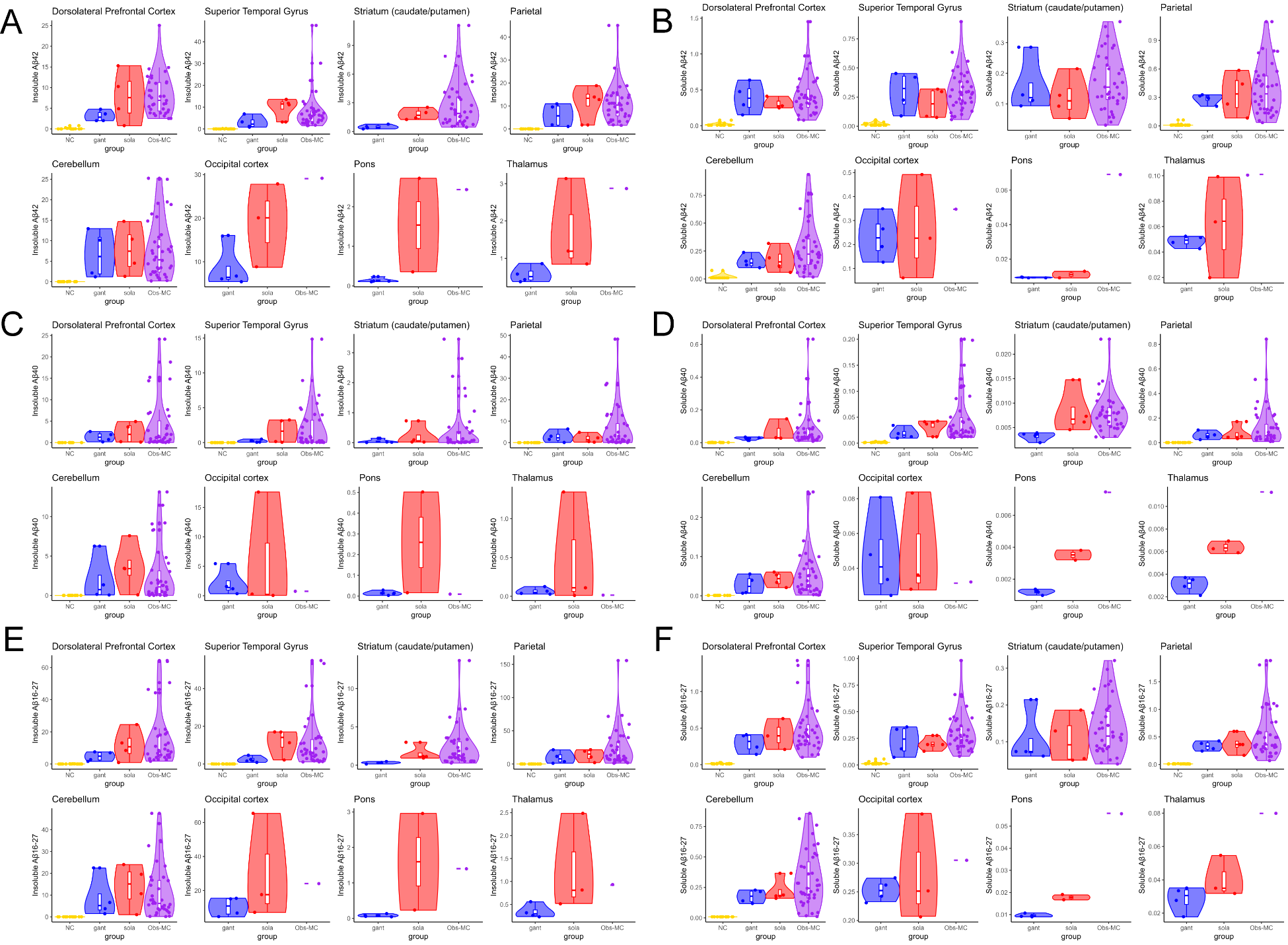

**Supplementary Figure S3.** Scatter plots of quantitative measures of the (A) insoluble Aβ_1-15_ (ng/mg brain), (B) insoluble Aβ_3pGlu-15_ (ng/mg brain), (C) soluble Aβ_1-15_ (ng/mg brain) and (D) Aβ_3pGlu-15_ (ng/mg brain) in all the different brain regions (dorsolateral prefrontal cortex, superior temporal gyrus, striatum, parietal cortex, cerebellum, occipital cortex, pons and thalamus) from the postmortem brains of age-relevant, cognitively normal non-DIAD controls, DIAD mutation carriers treated with gantenerumab or solanezumab and untreated observational DIAD mutation carriers.
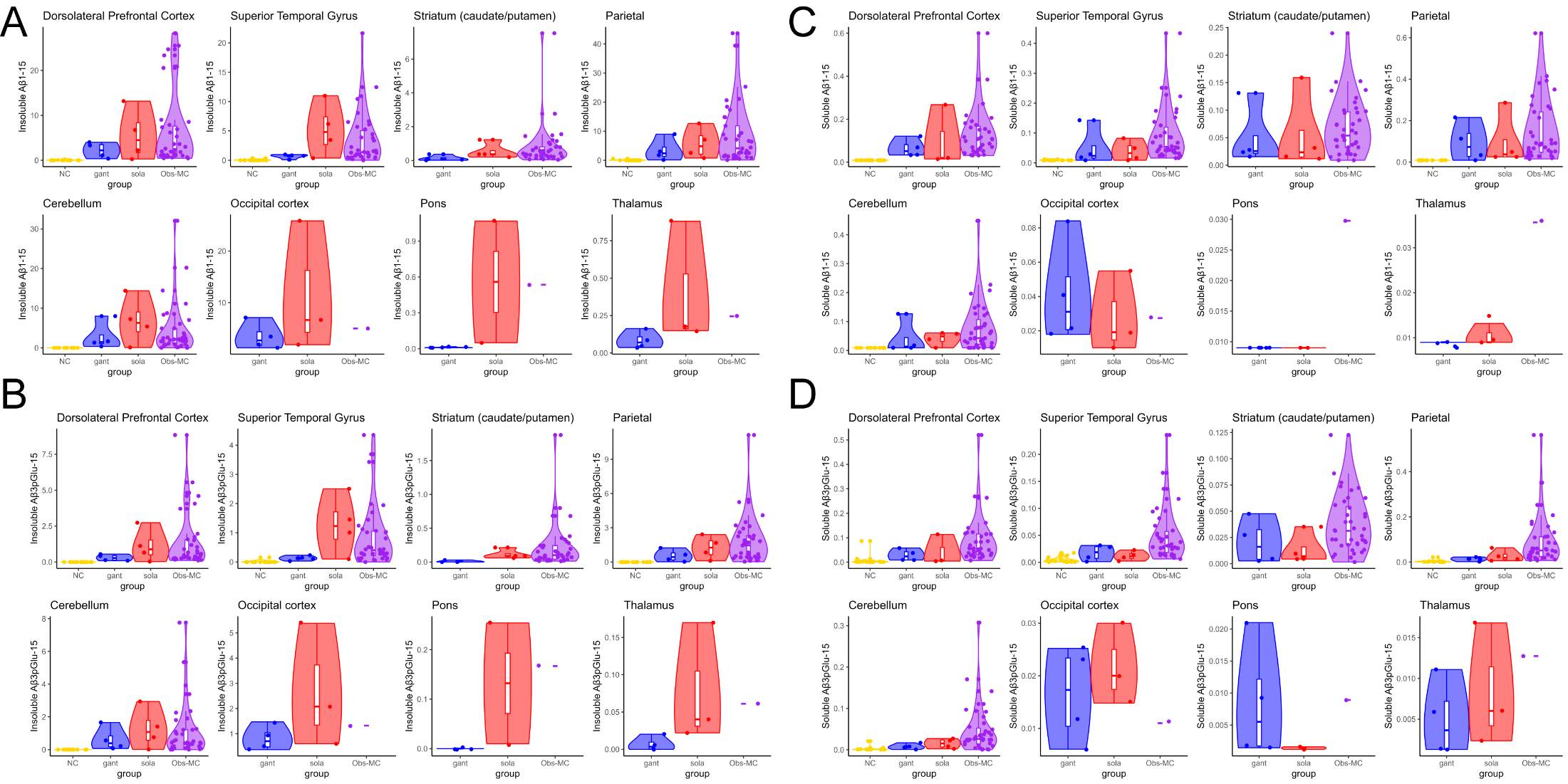

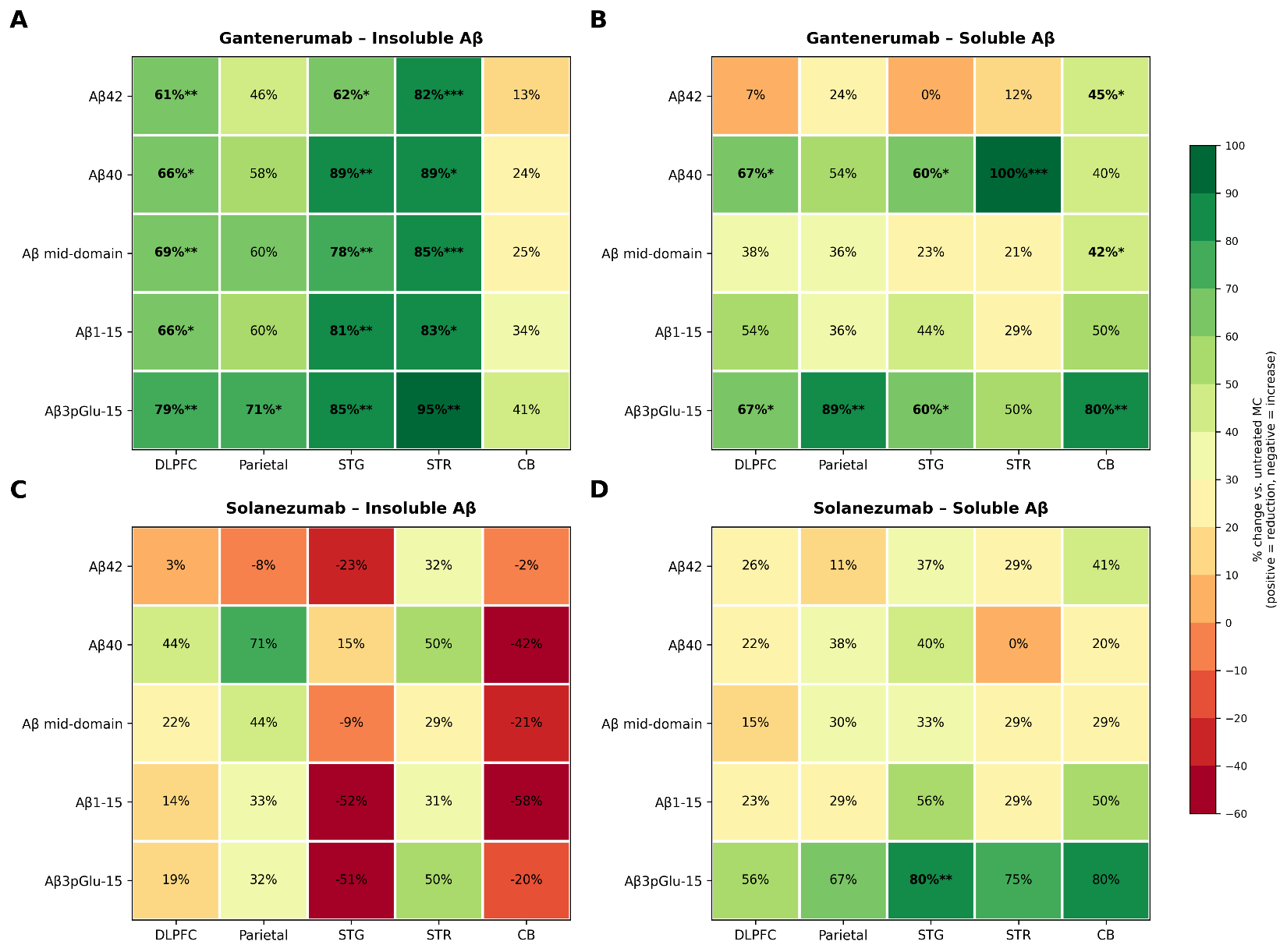
**Supplementary Figure S4.** Heatmap depicting the percent change in brain Aβ proteoform levels (ng/mg brain) in the treated DIAN-TU participants treated with gantenerumab or solanezumab relative to untreated DIAN Obs MC participants, across different brain regions (DLPFC, dorsolateral frontal cortex; parietal, STG, superior temporal cortex; STR, striatum; CB, cerebellum). Gantenerumab (A) insoluble and (B) soluble fractions. Solanezumab (C) insoluble and (D) soluble fractions. Cell color and value indicate percent change relative to untreated DIAN Obs-MC (positive values/green = reduction; negative values/red = increase), using a discrete 10% color scale. Group comparisons were performed using Welch's two-sample t-test, with p-values corrected for multiple comparisons within each treatment arm using the Benjamini-Hochberg false discovery rate (FDR) procedure. Asterisks denote FDR-corrected significance: *p<0.05, **p<0.01, ***p<0.001.

**Supplementary Figure S5.** Aβ42 (ng/mg brain) levels and gantenerumab dose response. Gantenerumab dose response at postmortem using high resolution mass spectrometry from all the DIAN-TU-001 gantenerumab (n=4) arm participants (blue) compared to the non-drug treated DIAN Obs-MC (black).

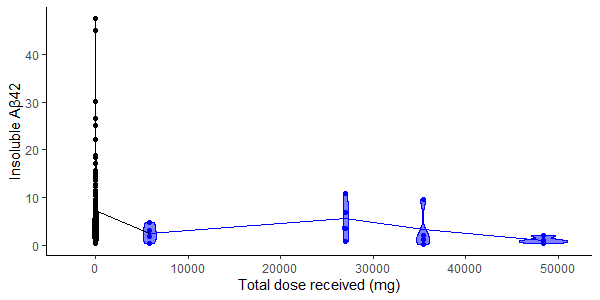

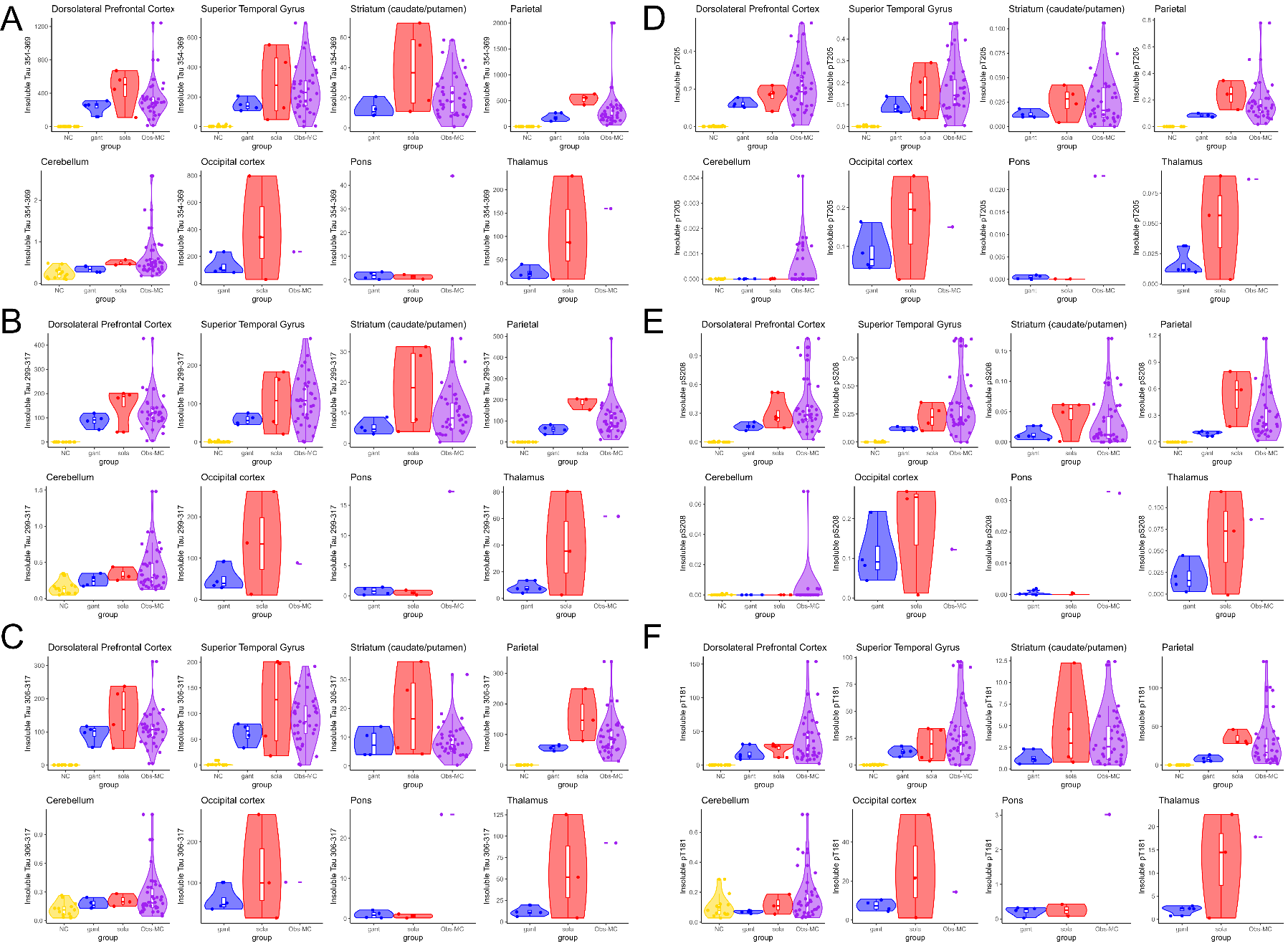
**Supplementary Figure S6.** Scatter plots of quantitative measures of the (A) insoluble MTBR-tau354-369 (ng/mg brain), (B) MTBR-tau299-317 (ng/mg brain), (C) MTBR-tau306-317 (ng/mg brain), (D) pT205 (ng/mg brain), (E) pS208 (ng/mg brain) and (F) pT181 (ng/mg brain) in all the different brain regions (dorsolateral prefrontal cortex, superior temporal gyrus, striatum, parietal cortex, cerebellum, occipital cortex, pons and thalamus) from the postmortem brains of age-relevant, cognitively normal non-DIAD controls, DIAD mutation carriers treated with gantenerumab or solanezumab and untreated observational DIAD mutation carriers.

**Supplementary Figure S7.** Heatmap depicting the percent change in insoluble brain tissue tau species, three microtubule-binding-region (MTBR) fragments (MTBR-tau354-369; MTBR-tau306-317 (3R) and MTBR-tau299-317(4R)) and five phosphorylated tau species (pT181, pS202, pT205, pS208 and pT217), in treated DIAN-TU participants relative to untreated DIAN Obs-MC controls, across five brain regions (DLPFC, dorsolateral prefrontal cortex; Parietal; STG, superior temporal gyrus; STR, striatum; CB, cerebellum). (A) Insoluble and (B) soluble tau Gantenerumab. (C) Insoluble and (D) soluble tau Solanezumab. Cell color and value indicate percent change relative to untreated DIAN Obs-MC (positive values/green = reduction; negative values/red = increase), using a discrete 10% color scale. Group comparisons were performed using Welch's two-sample t-test, with p-values corrected for multiple comparisons within each treatment arm using the Benjamini-Hochberg false discovery rate (FDR) procedure. Asterisks denote FDR-corrected significance: *p<0.05, **p<0.01, ***p<0.001. "NA" indicates a comparison that could not be estimated (zero variance in both groups).

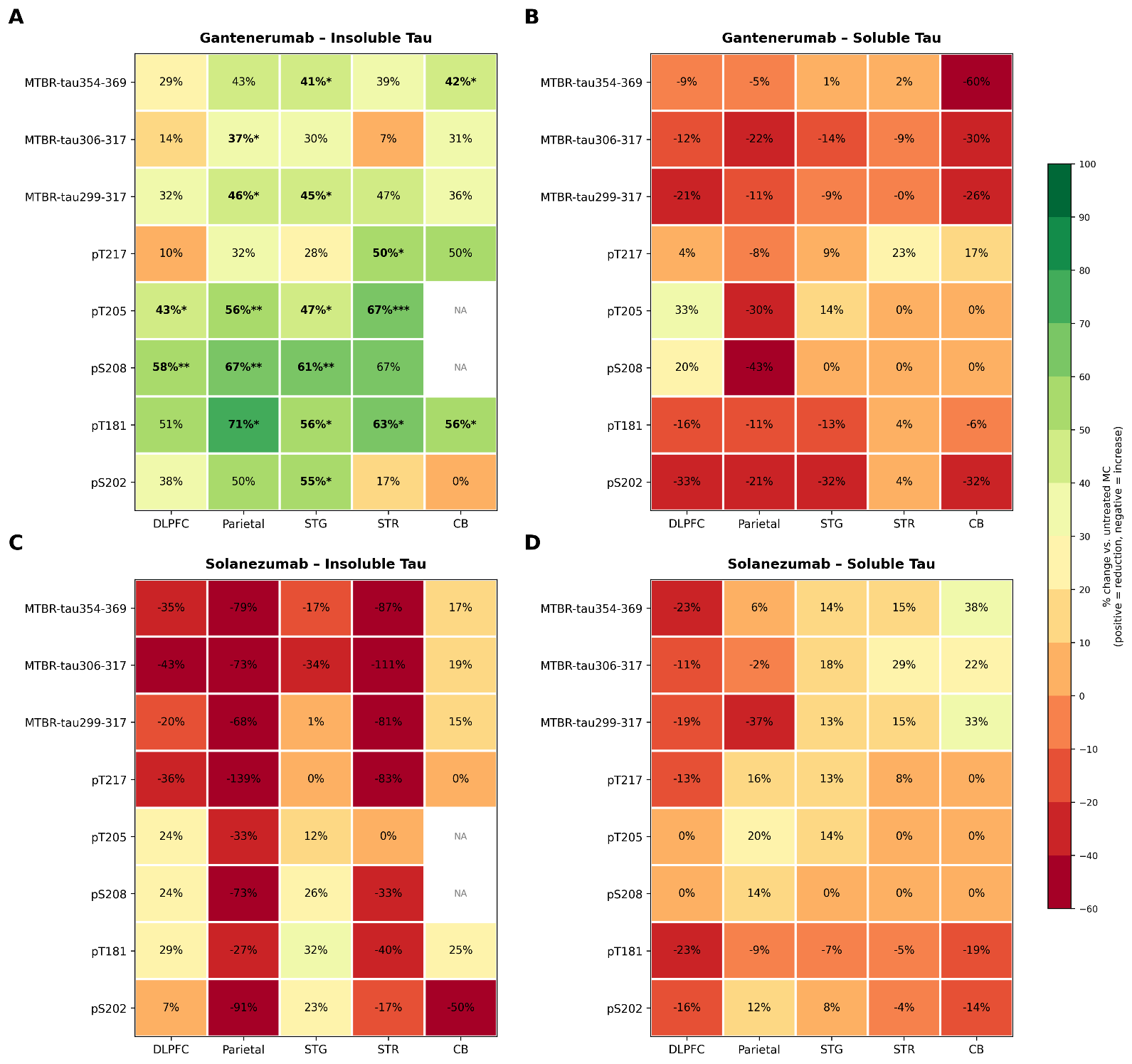

**Supplementary Figure S8**. Scatter plots of quantitative estimation of soluble (ng/mg brain) (A) MTBR-tau354-369, (B) MTBR-tau306-317 (3R), (C) MTBR-tau299-317 (4R) and phosphorylated tau levels (ng/mg brain) (D) insoluble and (E) soluble pT217, soluble (F) pT205, (G) pS208, (H) pT181, and (I) insoluble and (J) soluble pS202 in different brain regions of postmortem brains (dorsolateral prefrontal cortex, superior temporal gyrus, striatum, parietal and cerebellum) the postmortem brains of age-relevant, cognitively normal non-DIAD controls, DIAD mutation carriers treated with gantenerumab or solanezumab and untreated observational DIAD mutation carriers. The total cumulative dose (mg) of the drug received in the clinical trial for gant (blue) and sola (red) is highlighted in color gradient for each participant.

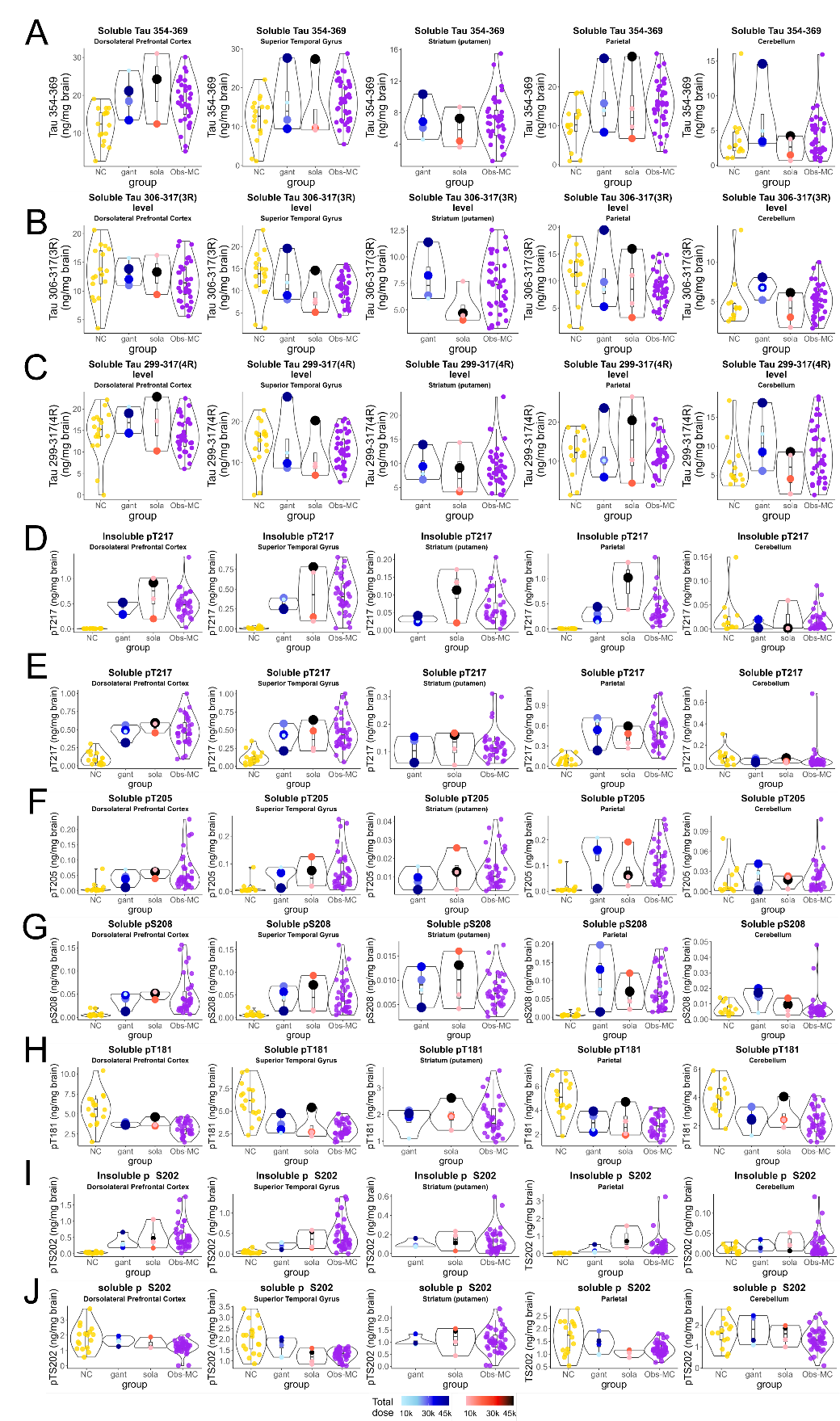

**Supplementary Table 1.** Participant characteristics of the non-DIAD controls and neuropathological demographics at postmortem from Goizueta Alzheimer’s disease research center at Emory University.

| Sample ID | Braak Stage | CERAD | Thal | PMI (hours) | Age at Death (years) | *ApoE* genotype | Race | Sex |
| --- | --- | --- | --- | --- | --- | --- | --- | --- |
| 1 | 0 | 0 | 0 | 25.5 | 31-36 | ε2ε4 | w | m |
| 2 | 0 | 0 | 0 | 22.5 | 30-35 | ε3ε3 | w | f |
| 3 | 0 | 0 | 2 | 31 | 40-45 | ε3ε4 | w | m |
| 4 | 0 | 0 | 0 | 15 | 41-46 | ε3ε3 | b | f |
| 5 | 0 | 0 | 0 | 28 | 42-47 | ε3ε3 | w | f |
| 6 | 0 | 0 | 0 | 6.5 | 46-50 | ε3ε3 | w | f |
| 7 | I | 0 | 0 | 22.5 | 50-55 | ε3ε3 | w | m |
| 8 | 0 | 0 | 2 | 3 | 51-56 | ε3ε4 | w | f |
| 9 | II | 0 | 0 | 6.5 | 49-53 | ε4ε4 | b | m |
| 10 | I | 0 | 0 | 12.5 | 55-60 | N/A | w | m |
| 11 | I | 0 | 2 | 17 | 54-58 | ε3ε3 | b | f |
| 12 | II | 0 | 0 | 10 | 56-60 | ε3ε3 | w | m |
| 13 | N/A | N/A | N/A | 6 | 58-63 | ε3ε3 | w | m |
| 14 | I | 0 | 0 | 6 | 57-62 | ε2ε3 | b | m |
| 15 | I | 0 | 1 | 8 | 60-65 | ε3ε4 | b | f |
| 16 | II | 0 | 2 | <12 | 58-63 | ε3ε4 | b | m |
| 17 | II | 0 | 1 | 6 | 61-66 | N/A | b | f |
| 18 | I | 1 | 1 | <12 | 65-70 | ε3ε3 | w | m |

*APOE,* Apolipoprotein E genotype; m, male; f, female, w, white; b, black; PMI; postmortem interval, Thal phases: Aβ/amyloid plaque score (0-5); Braak stage: neurofibrillary tangle score (0-VI) CERAD: Neuritic plaque score; N/A, not available

**Supplementary Table 2.** Results of the analysis of dose-dependence for gantenerumab treatment for the different Aβ peptides.

| Region |  | Estimate | Standard Error | DF | t-Value | Pr > \|t\| |
| --- | --- | --- | --- | --- | --- | --- |
| Insoluble Aβ42 | Intercept | 7.4001 | 0.7455 | 41.1 | 9.93 | <0.0001 |
|  | Total Dose | -0.00012 | 0.000073 | 38.9 | -1.66 | 0.11 |
| Insoluble Aβ1-15 | Intercept | 4.7107 | 0.8086 | 42.6 | 5.83 | <0.0001 |
|  | Total Dose | -0.00009 | 0.00008 | 40.4 | -1.07 | 0.29 |
| Insoluble Aβ40 | Intercept | 3.1222 | 0.7019 | 42.2 | 4.45 | <0.0001 |
|  | Total Dose | -0.00005 | 0.000069 | 40.1 | -0.78 | 0.44 |
| Insoluble Aβ16-27 | Intercept | 12.7116 | 2.044 | 42.7 | 6.22 | <0.0001 |
|  | Total Dose | -0.00023 | 0.000201 | 40.4 | -1.15 | 0.26 |
| Insoluble Aβ3pGlu-15 | Intercept | 1.0492 | 0.1713 | 42.9 | 6.12 | <0.0001 |
|  | Total Dose | -0.00002 | 0.000017 | 40.6 | -1.29 | 0.20 |

**Supplementary Table 3**. Results of the mixed model analysis for the postmortem cohort investigating the group changes of the respective biomarkers in different brain regions for each drug.

|  | Brain Region | Estimate | SE | DF | t Value | Pr > \|t\| |
| --- | --- | --- | --- | --- | --- | --- |
| Insoluble tau354-369 | | | | | | |
| Gantenerumab vs Obs-MC | CB | 0.536 | 83.071 | 217 | 0.01 | 0.9949 |
| Solanezumab vs Obs-MC | CB | -36.6123 | 92.9836 | 217 | -0.39 | 0.6942 |
| Gantenerumab vs Obs-MC | DLPFC | -89.837 | 83.4704 | 217 | -1.08 | 0.283 |
| Solanezumab vs Obs-MC | DLPFC | 120.6 | 83.4704 | 217 | 1.44 | 0.1499 |
| Gantenerumab vs Obs-MC | Parietal | -128.14 | 83.3669 | 217 | -1.54 | 0.1257 |
| Solanezumab vs Obs-MC | Parietal | 198.04 | 93.2481 | 217 | 2.12 | **0.0348** |
| Gantenerumab vs Obs-MC | STG | -98.9917 | 83.1669 | 217 | -1.19 | 0.2352 |
| Solanezumab vs Obs-MC | STG | 43.1979 | 83.1669 | 217 | 0.52 | 0.604 |
| Gantenerumab vs Obs-MC | STR(Pu) | -6.7453 | 83.0754 | 217 | -0.08 | 0.9354 |
| Solanezumab vs Obs-MC | STR(Pu) | 19.1142 | 83.0754 | 217 | 0.23 | 0.8182 |
| Insoluble tau299-317 (4R) | | | | | | |
| Gantenerumab vs Obs-MC | CB | 0.1871 | 25.2449 | 217 | 0.01 | 0.9941 |
| Solanezumab vs Obs-MC | CB | -13.7162 | 28.1145 | 217 | -0.49 | 0.6261 |
| Gantenerumab vs Obs-MC | DLPFC | -38.9191 | 25.3607 | 217 | -1.53 | 0.1263 |
| Solanezumab vs Obs-MC | DLPFC | 27.1714 | 25.3607 | 217 | 1.07 | 0.2852 |
| Gantenerumab vs Obs-MC | Parietal | -51.0436 | 25.331 | 217 | -2.02 | **0.0451** |
| Solanezumab vs Obs-MC | Parietal | 60.9833 | 28.1919 | 217 | 2.16 | **0.0316** |
| Gantenerumab vs Obs-MC | STG | -46.7752 | 25.2729 | 217 | -1.85 | 0.0656 |
| Solanezumab vs Obs-MC | STG | -0.3594 | 25.2729 | 217 | -0.01 | 0.9887 |
| Gantenerumab vs Obs-MC | STR(Pu) | -4.1752 | 25.2464 | 217 | -0.17 | 0.8688 |
| Solanezumab vs Obs-MC | STR(Pu) | 8.5592 | 25.2464 | 217 | 0.34 | 0.7349 |
| Insoluble tau306-317 (3R) | | | | | | |
| Gantenerumab vs Obs-MC | CB | -0.03661 | 19.9537 | 217 | 0 | 0.9985 |
| Solanezumab vs Obs-MC | CB | -13.2234 | 22.2797 | 217 | -0.59 | 0.5535 |
| Gantenerumab vs Obs-MC | DLPFC | -13.3838 | 20.0475 | 217 | -0.67 | 0.5051 |
| Solanezumab vs Obs-MC | DLPFC | 48.8345 | 20.0475 | 217 | 2.44 | **0.0157** |
| Gantenerumab vs Obs-MC | Parietal | -33.8298 | 20.0233 | 217 | -1.69 | 0.0926 |
| Solanezumab vs Obs-MC | Parietal | 53.6813 | 22.3421 | 217 | 2.4 | **0.0171** |
| Gantenerumab vs Obs-MC | STG | -26.2868 | 19.9763 | 217 | -1.32 | 0.1896 |
| Solanezumab vs Obs-MC | STG | 30.5837 | 19.9763 | 217 | 1.53 | 0.1272 |
| Gantenerumab vs Obs-MC | STR(Pu) | -0.23 | 19.9548 | 217 | -0.01 | 0.9908 |
| Solanezumab vs Obs-MC | STR(Pu) | 9.9872 | 19.9548 | 217 | 0.5 | 0.6172 |

**Supplementary Table 4**. Quantitative measures (ng/mg brain) of the sarkosyl insoluble and soluble phospho tau proteoforms from the different brain regions investigated from NC, DIAN Obs-MC and DIAN-TU drug treated patients. All data are represented as mean (± standard deviation).

|  | **Brain region** | **DIAN Obs-MC** | **NC** | **Gantenerumab (n =4)** | **Gantenerumab vs DIAN Obs MC *p*-values** | **Solanezumab (n =4)** | **Solanezumab vs DIAN Obs MC *p*-values** |
| --- | --- | --- | --- | --- | --- | --- | --- |
| Insoluble Tau | | | | | | |  |
| pS202 | CB | 0.02 ± 0.03 (n=38) | 0.01 ± 0.01 (n=13) | 0.02 ± 0.01 (n=4) | 0.89 | 0.03 ± 0.02 (n=3) | 0.68 |
|  | DLPFC | 0.56 ± 0.41 (n=34) | 0.03 ± 0.02 (n=17) | 0.35 ± 0.21 (n=4) | 0.15 | 0.52 ± 0.38 (n=4) | 0.86 |
|  | Parietal | 0.46 ± 0.57 (n=35) | 0.03 ± 0.01 (n=16) | 0.23 ± 0.2 (n=4) | 0.13 | 0.88 ± 0.64 (n=3) | 0.37 |
|  | STG | 0.47 ± 0.38 (n=37) | 0.05 ± 0.04 (n=17) | 0.21 ± 0.08 (n=4) | **0.001** | 0.36 ± 0.24 (n=4) | 0.43 |
|  | STR(Pu) | 0.12 ± 0.13 (n=38) | *NA* | 0.1 ± 0.04 (n=4) | 0.50 | 0.14 ± 0.09 (n=4) | 0.69 |
| pT217 | CB | 0.02 ± 0.02 (n=38) | 0.02 ± 0.04 (n=13) | 0.01 ± 0.01 (n=4) | 0.13 | 0.02 ± 0.03 (n=3) | 0.82 |
|  | DLPFC | 0.5 ± 0.25 (n=34) | 0.01 ± 0 (n=17) | 0.45 ± 0.11 (n=4) | 0.57 | 0.68 ± 0.36 (n=4) | 0.39 |
|  | Parietal | 0.38 ± 0.27 (n=35) | 0.01 ± 0 (n=16) | 0.26 ± 0.13 (n=4) | 0.19 | 0.91 ± 0.48 (n=3) | 0.19 |
|  | STG | 0.43 ± 0.23 (n=37) | 0.01 ± 0.01 (n=17) | 0.31 ± 0.08 (n=4) | 0.06 | 0.43 ± 0.36 (n=4) | 0.97 |
|  | STR(Pu) | 0.06 ± 0.04 (n=38) | *NA* | 0.03 ± 0.01 (n=4) | **<0.001** | 0.11 ± 0.06 (n=4) | 0.24 |
| Soluble tau | | | | | | | |
| pS202 | CB | 1.36 ± 0.47 (n=39) | 1.6 ± 0.64 (n=13) | 1.8 ± 0.7 (n=4) | 0.30 | 1.55 ± 0.47 (n=4) | 0.48 |
|  | DLPFC | 1.23 ± 0.41 (n=35) | 1.86 ± 0.79 (n=17) | 1.64 ± 0.29 (n=4) | 0.06 | 1.43 ± 0.39 (n=3) | 0.47 |
|  | Parietal | 1.2 ± 0.25 (n=37) | 1.67 ± 0.65 (n=16) | 1.45 ± 0.38 (n=4) | 0.28 | 1.06 ± 0.14 (n=4) | 0.15 |
|  | STG | 1.3 ± 0.24 (n=38) | 2.03 ± 0.71 (n=17) | 1.72 ± 0.4 (n=4) | 0.12 | 1.2 ± 0.35 (n=4) | 0.60 |
|  | STR(Pu) | 1.07 ± 0.44 (n=39) | *NA* | 1.03 ± 0.2 (n=4) | 0.81 | 1.11 ± 0.51 (n=4) | 0.87 |
| pT217 | CB | 0.06 ± 0.11 (n=39) | 0.02 ± 0.04 (n=13) | 0.05 ± 0.03 (n=4) | 0.54 | 0.06 ± 0.02 (n=4) | 0.66 |
|  | DLPFC | 0.48 ± 0.2 (n=35) | 0.01 ± 0 (n=17) | 0.46 ± 0.1 (n=4) | 0.73 | 0.54 ± 0.08 (n=3) | 0.33 |
|  | Parietal | 0.5 ± 0.22 (n=37) | 0.01 ± 0 (n=16) | 0.54 ± 0.22 (n=4) | 0.75 | 0.42 ± 0.14 (n=4) | 0.40 |
|  | STG | 0.46 ± 0.21 (n=38) | 0.01 ± 0.01 (n=17) | 0.42 ± 0.16 (n=4) | 0.62 | 0.4 ± 0.2 (n=4) | 0.58 |
|  | STR(Pu) | 0.13 ± 0.06 (n=39) | *NA* | 0.1 ± 0.05 (n=4) | 0.47 | 0.12 ± 0.05 (n=4) | 0.90 |
| pT205 | CB | 0.02 ± 0.02 (n=39) | 0.02 ± 0.02 (n=13) | 0.02 ± 0.02 (n=4) | 0.97 | 0.02 ± 0.01 (n=4) | 0.58 |
|  | DLPFC | 0.06 ± 0.05 (n=35) | 0.01 ± 0.02 (n=17) | 0.04 ± 0.02 (n=4) | 0.28 | 0.06 ± 0.01 (n=3) | 0.86 |
|  | Parietal | 0.1 ± 0.07 (n=37) | 0.01 ± 0.03 (n=16) | 0.13 ± 0.09 (n=4) | 0.56 | 0.08 ± 0.08 (n=4) | 0.61 |
|  | STG | 0.07 ± 0.07 (n=38) | 0.01 ± 0.02 (n=17) | 0.06 ± 0.03 (n=4) | 0.49 | 0.06 ± 0.05 (n=4) | 0.68 |
|  | STR(Pu) | 0.01 ± 0.01 (n=39) | *NA* | 0.01 ± 0.01 (n=4) | 0.20 | 0.01 ± 0.01 (n=4) | 0.98 |
| pS208 | CB | 0.01 ± 0.01 (n=39) | 0.01 ± 0 | 0.01 ± 0.01 (n=4) | 0.15 | 0.01 ± 0 (n=4) | 0.85 |
|  | DLPFC | 0.05 ± 0.04 (n=35) | 0.01 ± 0.01 | 0.04 ± 0.02 (n=4) | 0.43 | 0.05 ± 0.01 (n=3) | 0.87 |
|  | Parietal | 0.07 ± 0.05 (n=37) | 0.01 ± 0 | 0.1 ± 0.08 (n=4) | 0.44 | 0.06 ± 0.04 (n=4) | 0.81 |
|  | STG | 0.05 ± 0.04 (n=37) | 0.01 ± 0 | 0.05 ± 0.02 (n=4) | 0.96 | 0.05 ± 0.04 (n=4) | 0.92 |
|  | STR(Pu) | 0.01 ± 0 (n=39) | *NA* | 0.01 ± 0 (n=4) | 0.58 | 0.01 ± 0.01 (n=4) | 0.45 |
| pT181 | CB | 2.26 ± 0.85 (n=39) | 0.1 ± 0.07 (n=13) | 2.39 ± 0.83 (n=4) | 0.78 | 2.7 ± 0.94 (n=4) | 0.43 |
|  | DLPFC | 3.18 ± 0.87 (n=35) | 0.23 ± 0.12(n=17) | 3.68 ± 0.27 (n=4) | **0.03** | 3.92 ± 0.62 (n=3) | 0.16 |
|  | Parietal | 2.71 ± 0.81 (n=36) | 0.27 ± 0.13(n=16) | 3.01 ± 0.85 (n=4) | 0.55 | 2.95 ± 1.29 (n=4) | 0.74 |
|  | STG | 3.04 ± 0.83 (n=38) | 0.38 ± 0.22(n=17) | 3.45 ± 0.95 (n=4) | 0.45 | 3.24 ± 1.46 (n=4) | 0.80 |
|  | STR(Pu) | 1.87 ± 0.64 (n=39) | *NA* | 1.8 ± 0.48 (n=4) | 0.81 | 1.97 ± 0.5 (n=4) | 0.72 |
| MTBR-tau354-369 | CB | 4.09 ± 2.86 (n=39) | 3.97 ± 3.91(n=13) | 6.54 ± 5.43 (n=4) | 0.44 | 2.54 ± 1.72 (n=4) | 0.17 |
|  | DLPFC | 18.34 ± 5.7 (n=35) | 12.21 ± 4.58(n=17) | 19.91 ± 5.46 (n=4) | 0.62 | 22.59 ± 9.39 (n=3) | 0.52 |
|  | Parietal | 15.58 ± 5.67 (n=37) | 10.55 ± 5.56(n=16) | 16.33 ± 7.96 (n=4) | 0.86 | 14.66 ± 9.3 (n=4) | 0.86 |
|  | STG | 16.42 ± 5.37 (n=38) | 12.01 ± 5.54(n=17) | 16.27 ± 8.09 (n=4) | 0.97 | 14.1 ± 8.86 (n=4) | 0.64 |
|  | STR(Pu) | 7.13 ± 2.86 (n=39) | *NA* | 6.98 ± 2.44 (n=4) | 0.92 | 6.03 ± 2.38 (n=4) | 0.44 |
| MTBR-tau306-317 (3R) | CB | 5.15 ± 1.93 (n=39) | 4.78 ± 3.03(n=13) | 6.68 ± 1.17 (n=4) | 0.07 | 4.04 ± 2.02 (n=4) | 0.36 |
|  | DLPFC | 11.72 ± 3.44 (n=35) | 13.22 ± 4.37(n=17) | 13.17 ± 2.09 (n=4) | 0.28 | 12.98 ± 3.43 (n=3) | 0.60 |
|  | Parietal | 8.81 ± 2.94 (n=37) | 10.92 ± 4.88(n=16) | 10.76 ± 6.11 (n=4) | 0.57 | 9.03 ± 5.65 (n=4) | 0.94 |
|  | STG | 10.62 ± 3.04 (n=38) | 13.3 ± 5.55(n=17) | 12.1 ± 5.22 (n=4) | 0.61 | 8.75 ± 4.08 (n=4) | 0.43 |
|  | STR(Pu) | 7.35 ± 2.55 (n=39) | *NA* | 8.03 ± 2.44 (n=4) | 0.63 | 5.23 ± 1.67 (n=4) | 0.08 |
| MTBR-tau299-317 (4R) | CB | 8.81 ± 4.55 (n=37) | 6.59 ± 4.09(n=13) | 11.06 ± 4.99 (n=4) | 0.44 | 5.88 ± 3.45 (n=4) | 0.19 |
|  | DLPFC | 14.15 ± 4.07 (n=35) | 14.37 ± 5.66(n=17) | 17.11 ± 3.09 (n=4) | 0.15 | 16.77 ± 6.31 (n=3) | 0.55 |
|  | Parietal | 11.28 ± 4.03 (n=36) | 12.14 ± 5.53(n=15) | 12.49 ± 7.65 (n=4) | 0.77 | 15.41 ± 9.9 (n=4) | 0.47 |
|  | STG | 13.08 ± 4 (n=38) | 14.57 ± 5.67(n=17) | 14.2 ± 8.08 (n=4) | 0.80 | 11.4 ± 6.02 (n=4) | 0.62 |
|  | STR(Pu) | 9.42 ± 4.04 (n=39) | *NA* | 9.46 ± 3.18 (n=4) | 0.98 | 8.05 ± 4.76 (n=4) | 0.61 |
